# Exploring support needs of people living with diabetes during the coronavirus COVID-19 pandemic: insights from a UK survey

**DOI:** 10.1101/2021.01.20.21249888

**Authors:** Sarah Sauchelli, Julia Bradley, Clare England, Aidan Searle, Alex Whitmarsh

## Abstract

**Background:** The coronavirus COVID-19 pandemic has radically compromised healthcare for people living with chronic conditions such as diabetes. Government-imposed restrictions to contain the spread of the virus has forced people to suddenly adjust their lifestyle. This study aimed to capture the impact of the pandemic on people living with diabetes and the views of these individuals on ways in which the information, advice and support they are receiving could be improved.

**Research design and methods:** An online anonymous survey was distributed across the UK during the first lockdown and initial easing. The survey comprised questions about confidence in diabetes self-management, resources used to obtain information, advice and support, and opinions on how these could be improved. Open-ended captured subjective experiences.

**Results:** The survey was completed by 773 adults living with diabetes (69.2% type 1, 28.5% type 2). There was notable variability in the impact of the pandemic on confidence in self-management, with confidence having deteriorated most commonly in the ability to take care of own mental wellbeing (37.0% respondents) and improved most commonly in maintaining a healthy weight (21.1% respondents). 41.2% of respondents living alone reported not receiving any outside support. The quality of information, advice and support received from the healthcare team was rated poorly by 37.2%. Respondents sought greater communication and tailored advice from their care team, clear and consistent information from the government and news channels, and improved understanding of diabetes and its challenges from their personal networks and employers.

**Conclusion:** Adjusting to the COVID-19 pandemic has strained the mental health and wellbeing of people living with diabetes. Diabetes care teams must receive assistance to support these individuals without risking further inequalities in access to healthcare. Equipping personal networks and employers with knowledge on diabetes and skills to support self-management may reduce the burden on the NHS.

**Significance of this study**

1. **What is already known about this subject?**
  - The COVID-19 pandemic has posed multiple challenges to the everyday life of people across the world.
  - People living with diabetes mellitus, particularly those with poor blood glucose are more vulnerable to developing the severe outcomes of COVID-19.
  - NHS prioritisation of COVID-19 has disrupted the availability of care for patients with chronic health conditions, including diabetes mellitus.
2. **What are the new findings?**
  - The pandemic generated a decrease in confidence in diabetes self-management, particularly regarding mental wellbeing (37.0%) and adhering to physical activity recommendations (32.0%) and a healthy eating pattern (29.6%). Greater access to the healthcare team and services, strategies to adjust self-care (with greater focus on mental health) and more external support are deemed as important to reinstate diabetes self-management.
  - Cancellation of appointments reduced patients’ access to knowledge on their glucose control and their confidence in diabetes self-management, it generated difficulties in switching between treatments and resulted in impoverished mental health and motivation to self-manage.
  - 41.2% of respondents living alone report not receiving support from outside their household.
  - Quality of information, advice and support received from the government and healthcare teams were perceived most poorly (% of respondents giving a rating of poor or very poor: 39.0% and 37.2% respectively). There is a request for greater transparency, higher quality information, improved contact, and an increased understanding of the condition by others.
3. **How might these results change the focus of research or clinical practice?**
  - There is a need to ensure equitable contact between healthcare teams and their patients, both for diabetes self-management and overall wellbeing.
  - A shift to remote consultations should include training practitioners to detect emotional distress in patients and the ability to refer patients to NHS or community-led mental health support.
  - A collective effort is needed to produce more stratified and consistent guidance, with clear messaging to minimise uncertainty and distress.
  - Further research and policy are needed to help patients identify a support network outside their direct care team and equip them with the knowledge and skills to provide adequate support.
  - Greater understanding on how some individuals were able to adjust their self-management successfully could assist care teams, relevant charities and policy makers to provide better support for those individuals who are struggling.

## INTRODUCTION

The coronavirus COVID-19 is a severe respiratory syndrome generated by infection by SARS coronavirus 2 (SARS-CoV-2). On the 30^th^ of January 2020, the WHO Emergency Committee declared COVID-19 a global health emergency[1], with approximately 41.77 million cases and 1.14 million deaths due to COVID-19 recorded worldwide within the first 10 months (https://ourworldindata.org/coronavirus). To contain the spread of the virus and protect the impact on the UK National Health Service (NHS), on the 23^d^ of March 2020 the UK government imposed a national lockdown and the prioritisation of COVID-19 patients across the NHS[2]. From June 1^st^ 2020, physical distancing measures restrictions have been imposed at varying degrees. Though these measures have been useful for flattening the rate of infection, they have caused severe disruption in the lives of people across the population[3,4], and in particular patient groups who rely on healthcare services[5].

For people living with diabetes, COVID-19 prioritisation in the NHS caused severe disruptions to healthcare provision available. Effects include the cancellation of routine check-up appointments (*e*.*g*. HbA1c and retinopathy checks), diabetes education sessions, and hospital services for non-urgent care. Additionally, support systems such as face-to-face peer support was suspended while digitally delivered solutions were accelerated[6]. As the pandemic persists, NHS England has published new guidelines encouraging a shift towards remote consultations whenever possible, the use of a case-by-case approach to evaluate the need for face-to-face reviews, and encouraging the uptake of digital self-management tools[7]. In addition to practical challenges in rolling out these guidelines across the NHS, the success of these changes in care delivery rely on patients’ ability to adapt and engage in technology-assisted self-care, as well as practitioners’ ability to interpret data from technology and their confidence in delivering care via remote consultations[8–10].

Given the nationally imposed restrictions and physical-distancing policies, and the limited access to healthcare teams, we expected the pandemic to have a notable impact on everyday diabetes management and the mental health of people living with diabetes, their parents, carers, and partners. This study aimed to capture this impact and the views of these individuals on how to improve the information, advice and support their receive during the pandemic.

## RESEARCH DESIGN AND METHODS

An online survey was developed by the NIHR Bristol Biomedical Research Centre in collaboration with the Diabetes UK South West team. The first draft of the survey was developed based on questions posted on the Diabetes UK forum, Facebook diabetes support groups, and discussions with diabetes support teams (*e*.*g*. Diabetes UK, Brigstowe) between the 1 ^st^ of April 2020 and 15 ^th^ of April 2020. The first draft was reviewed by Diabetes UK volunteers to ensure language, structure and question appropriateness.

The survey comprised a mixture of multiple-choice questions to quantify events and compare answers across groups, and open questions to gain insight on individual experiences and opinions. Responses were sought from people living with diabetes and their parents, carers and partners. Questions were adapted accordingly: parents, carers and partners were asked about their confidence in their ability to support diabetes self-management and their own experiences in obtaining information. The full survey, with all items and response options can be seen in Supplementary File 1.

### Outcome measures

- Demographic characteristics of the respondents, including diabetes type, postcode (first part only), age, gender, ethnicity, living situation.
- Information regarding the pandemic included physical distancing measures being taken at the time of completion (*e*.*g*. following stringent physical distancing or shielding), diagnosis of COVID-19 or presence of symptoms, and changes in living circumstances due to COVID-19.
- Confidence in diabetes self-management was rated (Likert scale 0-10) across several components of self-care, from ‘Could not do at all’ to 10 ‘Certain could do’.
- on a Likert scale (0-10) before and during the pandemic. They were asked the impact of appointment cancellation and what they thought would help ameliorate diabetes self-management.
- Information was gathered on the resources used for guidance on physical distancing measures, general diabetes self-management, and support for emotional wellbeing.
- Respondents provided ratings (five-point Likert scales) on ease of access to information and support regarding the various aspects of diabetes self-management (‘Very difficult’ to ‘very easy’), as well as the quality (‘Very poor’ to ‘Very good’) of the information, advice and support received from several sources (*e*.*g*. Government, Diabetes UK, Healthcare Team). When participants gave a ‘Very poor’ or ‘Poor’ rating, they were asked to provide their opinions on how to improve it.
- A final set of questions focused on the support received from respondents’ personal network.

The survey was distributed across the UK, between the 24 ^th^ of April 2020 and the 31 ^st^ of August 2020. A convenience sample of participants were recruited via dissemination of the survey by the networks of the NIHR Bristol BRC, the University of Bristol and Diabetes UK. Means of dissemination included research portals (*e*.*g*. the Oxford University Hospitals NHS Foundation Trust), social media (*e*.*g*. Facebook and Twitter), University of Bristol website, e-mail contacts and monthly newsletters (*e*.*g*. NIHR Bristol BRC, Diabetes UK). Participants were eligible for the study if they were aged 18 years or over, lived in the UK, and had either been diagnosed with diabetes or were the parent, carer, or partner of someone with diabetes.

Participants self-referred to the study by completing the survey and were not reimbursed for involvement. To ensure anonymity, participants were not asked to insert any identifiable personal information except for the first part of their postcode (to capture geographical area).

The data presented below reflects responses from people who identified themselves as living with diabetes. The number of respondents who were parents, carers, or partners of someone with diabetes was considered insufficiently large to draw conclusions. Results are nonetheless visible in Supplementary File 2.

### Analysis

Summary statistics show participant responses to survey questions. Results are presented for all participants with diabetes and by the main diabetes types. For questions on confidence in diabetes self-management, data are presented using medians and interquartile ranges. Differences in confidence scores before the pandemic and at survey completion were also calculated and participants were grouped by whether their scores decreased, were stable or increased.

Where multiple-choice questions included an ‘Other’ response, respondents were encouraged to expand on the answer. These were categorised by a single team researcher (JB) and agreement was sought with the principal investigator (SS). Where deemed more appropriate, a response was sorted into the pre-existing multiple-choice options (*e*.*g. “leaving the house only for exercise”* was classified as ‘adhering to physical/social distancing guidelines’). Open-ended questions were analysed using an inductive thematic approach. The first 15 responses of open-ended items were reviewed independently by two researchers (SS and JB) to generate an initial codebook for each item. The codebook was further refined following discussion with AS and CE until consensus was reached. Code names were renamed to reflect data and identify themes. This approach led to the development of a definitive coding framework by which all responses were coded. Analysis was carried out using the NVivo V.12 software package.

Ethical approval was obtained from the University of Bristol faculty research ethics committee (ref: 103163).

## RESULTS

A total of 773 people living with diabetes responded to the survey. (a further 79 participants were parents, partners, or carers of someone with diabetes). Though respondents were widely distributed across the UK, most came from the South East (n = 193) and South West (n = 142) regions of England.

### Demographic characteristics

Table 1 presents a breakdown of the demographic characteristics of respondents. Most were female (67.1%) and of white British ethnicity (90.1%). Mean age was of 47.9 (SD = 14.5, range 18 to 80) years. 69.2% of respondents reported living with type 1 diabetes mellitus (T1DM), 28.5% with type 2 diabetes mellitus (T2DM). Most respondents had not experienced symptoms of COVID-19 since the start of the pandemic (80.6%). The most common symptoms reported were coughing, shortness of breath, and fever. 66.8% of respondents were adhering to government social/physical distancing guidelines stipulated at the time of survey completion, 9.8% were voluntarily shielding despite not having received explicit instructions.

**Table 1.**
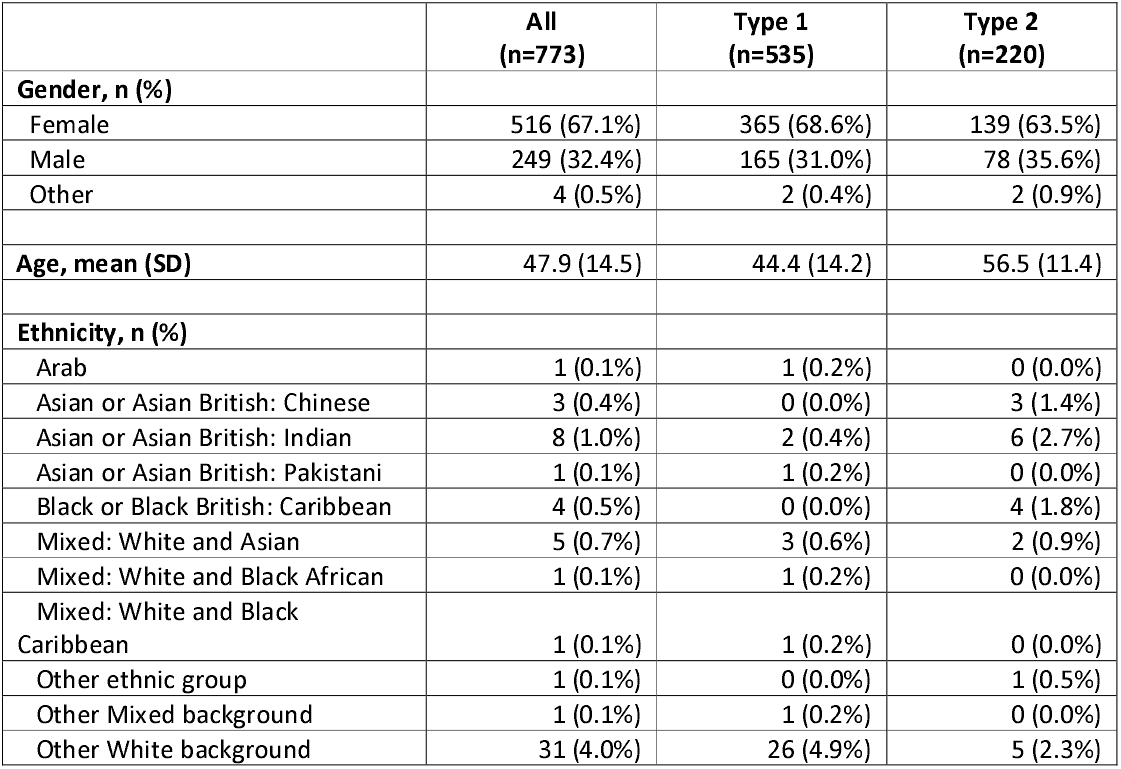
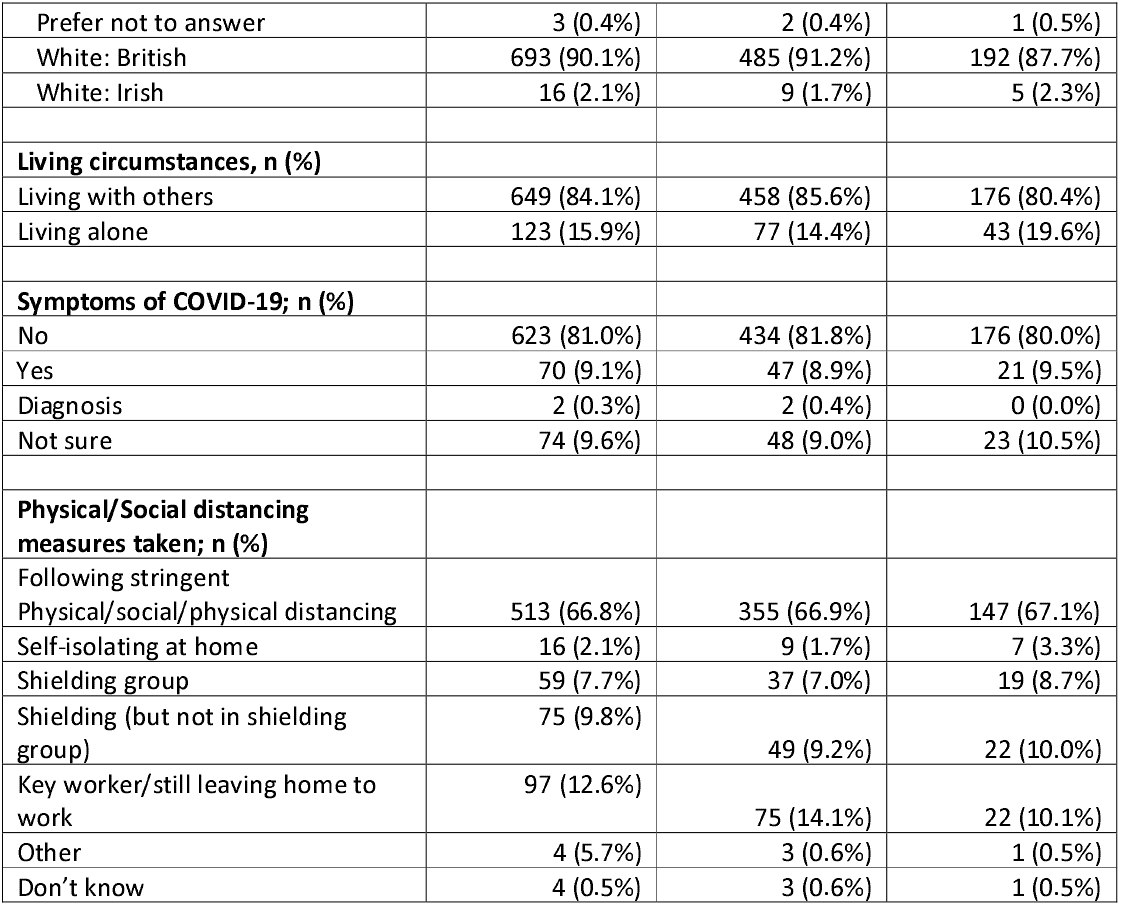
Demographic characteristics, COVID-19 symptoms and measures adopted by respondents with diabetes.

### Confidence in diabetes self-management

Confidence in self-management was impacted more notably in the lifestyle components of diabetes self-management (*e*.*g*. regular physical activity, healthy eating and maintenance of a healthy weight), and mental well-being (Figure 1). Change in confidence was mainly negative (poorer), particularly for mental well-being (37% showed a decrease), though a proportion of respondents displayed improvements.

**Figure 1:**
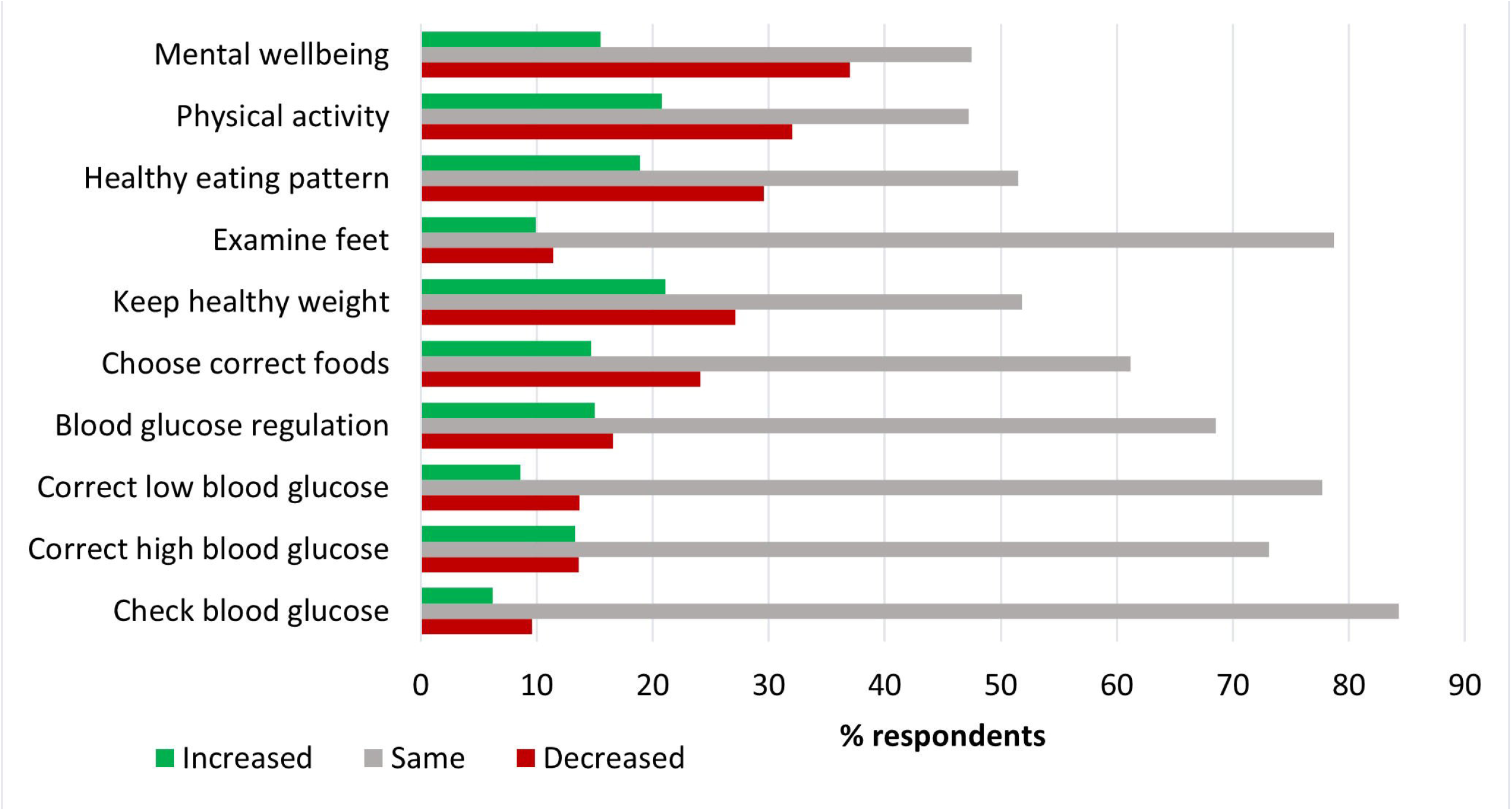
Change in confidence in diabetes self-management, at time of completion minus recall of confidence before the pandemic (n = 770). Positive score (green): increase. Negative score (red): decrease.

From the analysis, we identified three main themes on improving confidence in diabetes self-management: increased accessibility, adjusting self-care, and receipt of external support.

#### Accessibility

Respondents indicated needing greater access to their care team and the support provided for diabetes self-management, greater opportunities for physical activity, and easier access to the food they need to adhere to dietary recommendations:

*“Lockdown limited exercise which I rely on to control sugar levels. Readjustment of insulin due to my exercise is not straight forward”*

Several respondents indicated that receipt of blood glucose monitoring tools would have facilitated diabetes self-management:

*“Getting CGM on prescription”*

Further, access to clearer guidance on individual risk was deemed important to facilitate decision-making:

*“Preparation guides for how to manage sugar levels if you get coronavirus. Also guidelines on how to stay vigilant as a diabetic when carrying out daily activities”*

#### Adjusting self-care

Respondents described the need to adopt strategies that would help them adjust diabetes management. They were aware that unhealthy habits may be attributed to not having adapted to their new circumstances generated by the pandemic:

*“Dietary changes! Being less busy and at home more has meant a diet which is not so good for people with diabetes!”*

Respondents recognised the need to increase focus on mental health to reduce stress-induced glucose alterations:

*“My blood sugars have been more erratic due to the stress and worry for myself and my family, and they have been harder to keep under control*.”

Further, respondents recognised that this might require changes in doses or type of medication:

*“Reminders about changing insulin doses (via pump) in response to lower levels of physical activity*.*”*

#### External support

Need for assistance from personal network and wider community was deemed important to increase confidence. This included support from family and friends, greater adherence to physical distancing from others, and help in household tasks and childcare:

*“Lack of help with childcare means difficulty in exercising and more strain at home, so sugars are harder to look after*.*”*

Outside of these three factors, several respondents indicated that resumption of ‘normal’ life would be needed:

*“Once things get back to normal and I can get back to my routine”*

### Consequences of cancelled appointments

51.9% of respondents had at least one appointment cancelled at the time of survey completion. The impact was represented by four themes: lack of knowledge and confidence, difficulties in switching treatment, mental health, and empowerment in self-management:

#### Lack of knowledge and confidence

Cancellation of appointments resulted in uncertainty on glucose control, difficulties in interpreting information provided by monitoring devices, and lack of confidence in the actions to take to improve glucose control:

*“My self-confidence has plunged, and lack of follow-up hasn’t helped. The clinic cancelled appointments and I didn’t know who else to consult*.*”*

*“I have pretty much given up on self-management. I don’t know where I am in terms of my Hba1c so don’t know where I need to be heading*.*”*

#### Difficulties in switching treatment

Respondents indicated struggling to switch to other medications or changing doses and receiving adequate support to do so. They have had difficulties in using remote medical care, and experienced delayed or cancelled referrals to other services:

*“I was on a pathway of improving my treatment methods (a pump) but that has been paused”*

#### Mental health

Reduced support and advice regarding self-management or risk, and the cancellation of appointments was posing a strain on respondents’ mental health and motivation to continue self-management:

*“Although I don’t feel less able to self-manage, I have sometimes felt less motivated to manage my diabetes well. A result of general anxiety and poor sleep*.*”*

#### Empowerment in self-management

A few respondents indicated that they had managed to adapt to circumstances to improve self-management:

*“I have had to learn to cope and have read more and joined a Facebook diabetes support group, run by other diabetics”*

### Ease of access to information, advice and support

Respondents found it harder to receive support compared to information and advice. Access was more likely to be rated as ‘difficult’ or ‘very difficult’ in the domains ‘emotional wellbeing’ and ‘diabetes management if showing symptoms of COVID-19’ (Figure 2). There were clear differences between diabetes types in access to support: 42.5% of respondents with T2DM reported ‘difficult’ or ‘very difficult’ access to support for glucose control, compared to 28.9% of respondents with T1DM. Further details are in Supplementary File 2. Among those respondents who reported living alone, 41.2% indicated that they were not receiving support from outside the household. External support was received primarily from the family (68.7%), friends (67.2%) and neighbours (28.4%).

**Figure 2.**
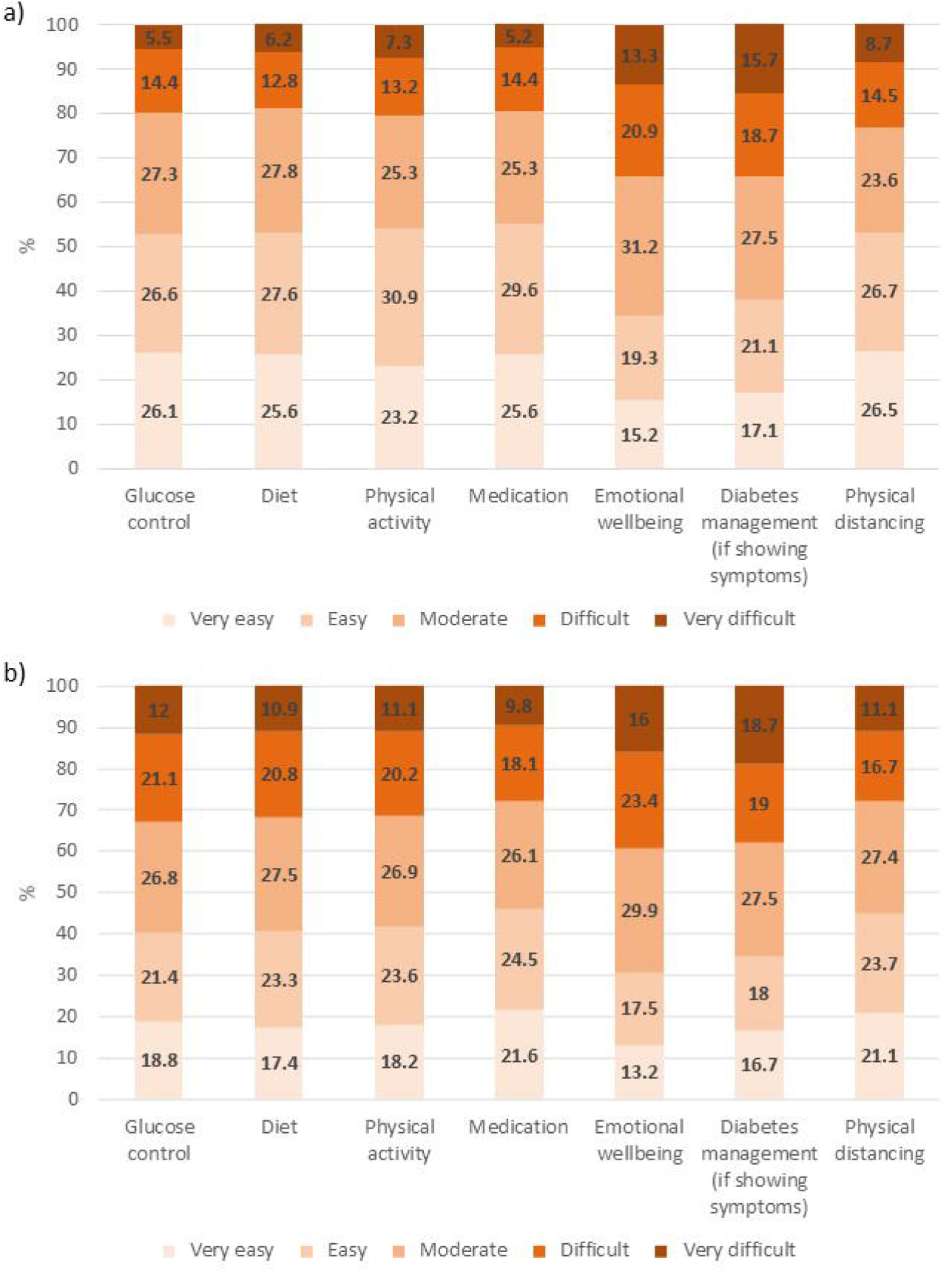
Rated difficulty in accessing a) information and advice, and b) support across diabetes self-management domains and adherence to physical distancing guidelines.

### Perceived quality of information, advice and support

Figure 3 shows respondents’ views on the quality of information, advice and support available across a wide range of sources. 39.0% of respondents rated the quality of government guidance and support as ‘poor’ or ‘very poor’, with lower scores from T1DM (41.8%) than T2DM (31.7%) (Supplementary File 2). Perceived quality in the guidance and support received from healthcare teams was similar, with 37% of respondents considering it as ‘poor’ or ‘very poor’. In this case ratings were poorer from T2DM (43.2%) compared to T1DM respondents (35.2%).

**Figure 3.**
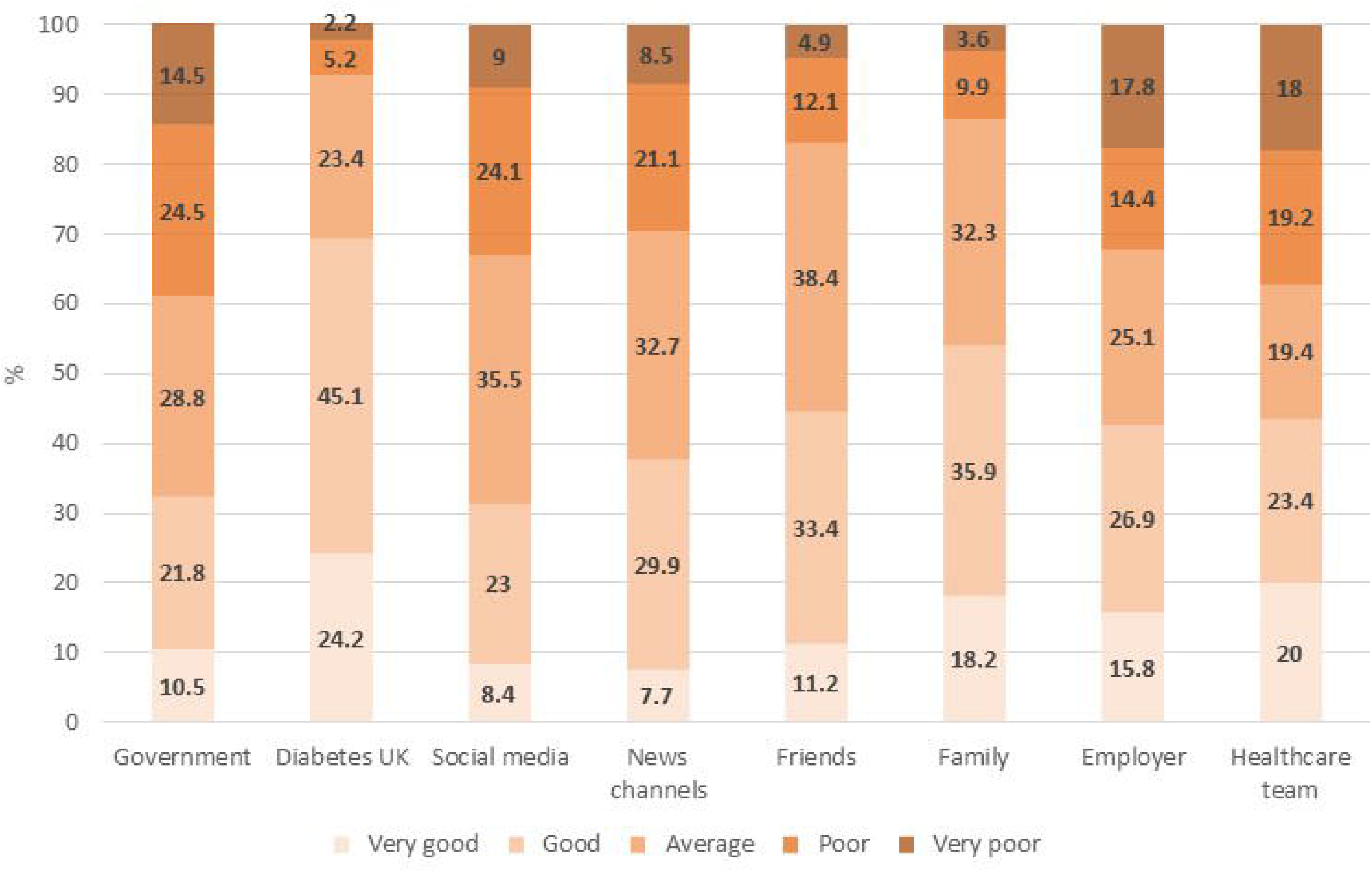
Reported quality of information, advice and support received from various resources.

Respondents who had provided ‘poor’ or ‘very poor’ scores were asked to suggest improvements that could be made. Figure 4 displays the main categories that emerged from analysis, subdivided according to source queried. From analysis, overarching themes were revealed: greater transparency, higher quality information and improved contact, and greater understanding of the condition.

**Figure 4.**
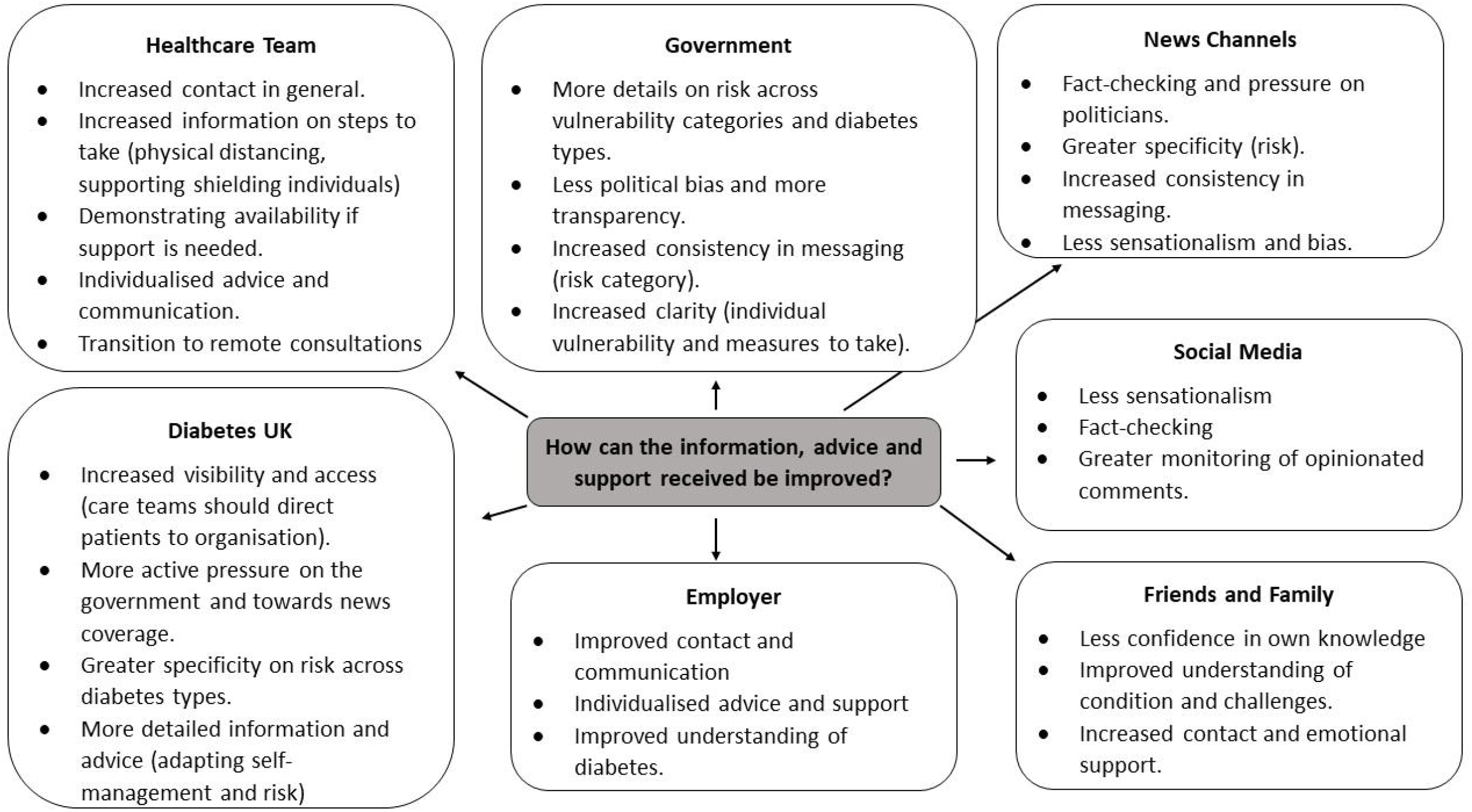
Main categories that emerged in respondents’ recommendations for improvement presented.

#### Greater transparency

Respondents expressed concerns regarding bias and tendency towards sensationalism in the information from the government, news channels and social media:

*“They over emphasise the negatives and cause fear or anxiety*.*”* (News Channels)

They requested these sources be more transparent in the evidence behind information and decision-making, greater fact-checking, objective reporting, and pressure on politicians to provide accurate information:

*“More challenge of government when information is inconsistent or ambiguous”* (48, T1DM, News Channels)

*“Fake news and anti-vac messaging to be removed promptly”* (Social Media)

*“It would be better if it came across as completely open and trustworthy*.*”* (Government)

#### Higher quality information

Respondents also communicated the need for improvement in information provided by healthcare teams, government, Diabetes UK, news channels and employers. They requested more information on precautionary measures to take in terms of shielding/physical distancing, how the personal network can help in emergencies, and diabetes self-management:

*“Needs more clarity for people like me who are “vulnerable” but have not received the NHS letter”* (Government)

*“Got told I had to return to work, no discussion about how worried that made me*.*”* (Employers)

*“When I had a hypo and was very mixed up and no one in the family intervened because of us being distanced inside the home*.*”* (Healthcare Team)

Data revealed that specificity was a frequent priority for improving the quality of information, distinguishing people with diabetes from other vulnerable people and differentiating between diabetes types. Greater specificity was sought for information on risk and for guidance on diabetes self-management:

*“Most of the dietary advice seems more geared to type 1 and doesn’t help me to lower my type 2 blood glucose”* (Diabetes UK)

*“Explain what the relevance of vulnerability to C-19 is in relation to what types of diabetics (type 1 or 2), those with complications etc, not just say ‘diabetics’“* (News Channels)

*“No specific policy for diabetics. Only general advice for people more vulnerable*.*”* (Employer)

Consistency in the information provided was also deemed important:

*“Changing risk category of Diabetes since the beginning. Caused lots of confusion*.*”* (Government)

Several respondents, however, communicated that they had noticed improvements with time:

*“The information was much more clear. Particularly as they spoke about T1 and T2 separately”* (Diabetes UK)

#### Improved contact and communication

Respondents frequently reported absence of their healthcare teams and employers, which had a negative impact on their mental health:

*“No contact from manager at this time and waiting for information has made this time more stressful*.*”* (Employer)

*“I have not received any information at all from my diabetes health care team”* (Healthcare Team)

There was a request for individualised contact and for the healthcare team to demonstrate availability if urgent support was needed:

*“I do feel that a quick phone call or more personal email would have been good*.*”* (Healthcare Team)

*“I have contacted my diabetic nurse several times, the only reply I have received is a text message suggesting I go to diabetes UK website”* (Healthcare Team)

*“Would be good to hear more of “please contact us if there is a problem” rather than always “stay away from the surgery*.*”* (Healthcare Team)

Opinions regarding the support provided by healthcare teams varied across respondents, as some indicated that their care team was responsive:

*“Rang me to check I was ok as check-up delayed. Could ring if I wanted to*.*”* (Healthcare Team)

Several respondents expressed an interest in remote consultations if this increased contact with their care team:

*“Improve access to diabetes team by telephone”* (Healthcare Team)

#### Increased understanding of diabetes

Respondents expressed wanting their personal networks and employers to have a better understanding of their condition and the challenges faced:

*“Unless you have an illness and keep being told about having a underlying illness is harmful during this time, you just don’t understand*.*”* (Friends and Family)

*“It would be good if they were a little better informed, particularly, now, about the increased risks posed to people with diabetes by Covid-19*.*”* (Employer)

This was important to enhance experienced support:

*“Friends are a very important source of general support”* (Friends and Family)

## CONCLUSIONS

This study provides valuable insight in the ways people living with diabetes have been impacted by the coronavirus COVID-19 pandemic. As expected, NHS prioritisation of COVID-19 has had a negative impact on the access and level of support most people with diabetes have during the pandemic, as experienced by people living with other chronic conditions[11]. Findings are consistent with growing evidence showing that government-imposed restrictions have had varied effects on people’s lifestyle and appraisal of COVID-19 guidance and support. Closure of sporting facilities and home confinement has contributed to a 28% increase in sitting time[4] and adoption of unhealthy dietary habits[12]. This reflects respondents’ decreased confidence in self-management in these domains. Yet, as found in this study, a proportion of individuals appear to have adapted successfully to pursue positive health-related behaviour change[12].

Findings resonate with research evidencing an increase in psychiatric disorders and diabetes-related emotional distress during COVID-19[13]. Diabetes increases vulnerability to psychiatric disorders, which can interfere with appropriate self-care and worsen diabetes prognosis[14]. Importantly, diabetes-related stress contributes to poor glucose control[15], which increases an vulnerability to the severity of COVID-19[16,17]. By capturing the experiences and opinions of people with diabetes, this study conveys actions that could be taken to ameliorate psychological distress.

Respondents called for increased contact and tailored guidance and support from their care teams. The reported poor quality of guidance and support received from the healthcare team may be due to respondents having higher expectations from their care team compared to other sources. Alternatively, it may be due the sudden change in access to the care team without patients having enough time to adjust their diabetes self-management. This is likely to have the greatest impact on patients who normally have more contact with their care team, those who rely on appointments to obtain insight on their glucose control, or those recently diagnosed. The response to remote consultations was generally positive, suggesting that it may be a feasible way forward. Health commissioners must ensure that care teams across the UK are adequately equipped and trained to ensure equal access to remote consultations. A practitioner’s familiarity with it is also key for its success, and previous work emphasises the importance of helping diabetes care teams recognise emotional distress in remote consultations[9].

This study shows that a collective effort should be made to improve the information provided to people with diabetes, focusing on stratified and consistent guidance on individual vulnerability, how to self-manage diabetes whilst minimising risk, and ensuring that people feel they can trust the entity communicating the information. Though greater communication and transparency has been greatly demanded throughout the pandemic[18], this study further shows how clear messaging is crucial to make vulnerable individuals feel safe in uncertain circumstances. It highlights how knowledge and confidence in self-management was heavily impacted by the cancellation of appointments, and extensive research and respondents’ commentaries from the survey demonstrate their importance for improving self-care in chronic conditions and mental wellbeing, even when under stress[19,20]. Given that the ability to adjust emerged as fundamental to improve diabetes self-management and mitigate the effects of cancelled appointments, communicating strategies to achieve this and providing support when needed may be highly beneficial. Examining how some individuals were able to adapt may be invaluable to inform stakeholders on which strategies are most effective.

Equipping the personal network with an increased understanding of diabetes and its challenges was also seen as important to increase the quality of support received. This aligns with extensive work demonstrating the value of a supportive immediate environment for the management of diabetes [20]. Accordingly, future research and action are needed to better understand why such a large proportion of respondents living alone were not receiving outside support. Isolation is a primary contributor to mental health difficulties[21], and identifying ways to connect people living alone with the community may be especially helpful during and after the pandemic.

Some methodological limitations need to be taken into consideration. The survey was distributed online. It may not accurately capture the views of individuals who engage less with healthcare teams or their community, and we did not reach people who are unable to access technology. Though multi-modal steps were taken to raise awareness of the survey, ethnic minorities and males were underrepresented. Alternative strategies should be adopted to target these groups, especially as the prevalence of diabetes is elevated in ethnic minority communities[22]. These limitations occurred in part due to the urgency of distributing the survey for Diabetes UK to take timely action, and obstacles faced due to the pandemic in engaging with key people that could facilitate wider participation. Despite its limitations, this study provides important insight into how the coronavirus COVID-19 pandemic has impacted people living with diabetes and their views on opportunities for improvement. As routine care is being cancelled due to increased infection rates and the roll out of vaccines, it is essential that experiences and opinions from the initial wave of the pandemic are incorporated in stakeholder decision-making.

## Data Availability

All anonymised data is available from the research team upon request.

https://cpb-eu-w2.wpmucdn.com/blogs.bristol.ac.uk/dist/3/495/files/2020/12/diabetes-survey-support-needs.pdf

## FUNDING

This work was supported by the Elizabeth Blackwell Institute, University of Bristol, the Wellcome Trust ISSF3 grant 204813/Z/16/Z. All authors are supported by the NIHR Biomedical Research Centre at University Hospitals of Bristol and Weston NHS Foundation Trust and the University of Bristol. The views expressed are those of the authors and not necessarily those of the NIHR or the Department of Health and Social Care.

## ACKNOWLEDGEMENTS

We thank Diabetes UK for their contribution to the design of the survey and dissemination via its networks. We thank research teams such as the Oxford Centre for Diabetes Endocrinology and Metabolism and the NIHR Oxford Biomedical Research Centre, Oxford University Hospitals NHS Foundation Trust for assisting in the distribution of the survey. Importantly, we thank the people living with diabetes and Diabetes UK volunteers who gave us feedback to ensure the survey was inclusive and appropriate for the diverse circumstances people living with diabetes may find themselves in during the COVID-19 pandemic.

## COMPETING INTERESTS

To enable Diabetes UK to take timely action from survey outcomes, three interim summary reports were produced for the Diabetes UK team, as well as a final one upon survey closure. Preparation of these reports did not impact the research project.

## Notes

### Competing Interest Statement

The authors have declared no competing interest.

### Author Declarations

University of Bristol faculty research ethics committee (ref: 103163).

