## Supplementary File 1 for "Exploring support needs of people living with diabetes during the coronavirus COVID-19 pandemic: insights from a UK survey"

**Participant Information Sheet**

A link to this sheet is provided in the first page of the online survey. This provides the respondent with further information on the purpose and their role in the study, confidentiality and use of data.

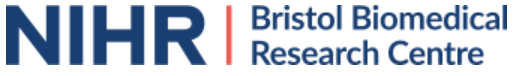

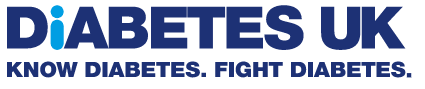

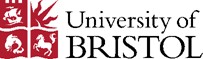

**PARTICIPANT INFORMATION SHEET**

**Survey title:** Identifying support needs of people with diabetes during the Coronavirus COVID-19 pandemic.

**Invitation paragraph**

We would like to invite you to take part in our research study. Before you decide whether to take part, it is important for you to understand why the research is being done and what it will involve. Please read through the following information carefully and discuss it with someone you trust if you wish. If there is anything that is unclear or you would like to receive more information, please do not hesitate to ask us by using the contact details provided.

**What is the purpose of the study?**

The purpose of this study is to help Diabetes UK to better understand and provide the type of support people with diabetes need during the coronavirus COVID-19 pandemic.

**Why have I been invited?**

You have been invited to take part in this study because you are over 18 years of age, have diabetes mellitus or a parent/carer/partner of someone with diabetes mellitus, and have expressed interest in taking part in this research by following the link on the study advertisement.

**Do I have to take part?**

It is up to you whether you would like to take part in this study. If you do decide to take part, you will be asked to give your consent by ticking the boxes on the web page. You are free to withdraw from the study at any time by exiting the survey and do not have to give a reason for doing so.

**What will happen to me if I take part and what will I have to do?**

Once you have read and understood the information about the study and have given your consent to take part on the survey page, by pressing Next you will be directed to the online survey. The survey will ask questions about a variety of topics related to the Coronavirus (COVID-19) outbreak. It should take around 15 minutes to complete. All the survey questions are optional. If you do not wish to answer a question, please leave it blank/do not press any of the response options and move on to the next question. The data collected during this survey will be anonymous.

**What are the possible disadvantages and risks of taking part?**

Taking part in this study is not expected to bring you any disadvantages. However, some of the survey questions will address issues that could be considered sensitive and responding to these questions may therefore cause some discomfort. For this reason, the survey questions are optional, allowing you to leave questions unanswered if you wish to avoid any sensitive topics.

**What are the possible benefits of taking part?**

Although we do not expect you to receive any immediate personal benefits from taking part in this study, the data collected could help improve the support provided to those with diabetes during the Coronavirus COVID-19 pandemic.

**Will my taking part in the study be kept confidential?**

All responses to the survey questions will be anonymous and not linked to any personal information that would identify you. The data collected as part of this study will shared with Diabetes UK to guide how they provide support to people with diabetes. It may also be shared with other researchers in the future as part of the collective response to the Coronavirus COVID-19. However, no-one will be able to trace your individual responses.

The sponsor of this study is the University of Bristol, based in the United Kingdom. The university, and specifically the NIHR Bristol Biomedical Research Centre will act as the data controller for this study. We are therefore responsible for looking after any information you provide and using it properly. If you withdraw from the study, we will keep the information that we have already obtained. The University of Bristol will keep your responses and identifiable information for 10 years after study completion in its secure, password-protected network. Subsequently, the information will be permanently removed in accordance with University of Bristol Policy.

Given the anonymity of the survey, it will be impossible for you to access your answers after completing the questionnaire. There is no route for us to trace your answers. As a university, we use identifiable information to conduct research to improve health, care and services. As a publicly funded organisation, we have to ensure that personally identifiable information about those who have agreed to take part in research is only utilised if it is in the public interest. This means that when you agree to complete the survey, we will use the demographic information you provide (e.g. gender, weight, height) in the ways needed to conduct and analyse the research.

**What will happen if I don’t want to carry on with the study?**

You are free to withdraw from the study at any time by exiting the survey web page. The answers you have already provided will be saved and may be used for analysis. However, these will be completely anonymous.

**What will happen to the results of the research study?**

The results of this study will be shared with Diabetes UK and published in peer-reviewed journals. All data included in publications will be anonymous, hence it will not be possible to identify you from the data. Results obtained will only be used by organisations and researchers to conduct research in accordance with the [UK Policy Framework for Health and Social Care Research](https://www.hra.nhs.uk/planning-and-improving-research/policies-standards-legislation/uk-policy-framework-health-social-care-research/).

**Who is organising and funding the research?**

This study is managed by Dr Sarah Sauchelli Toran, Dr Clare England, Dr Aidan Searle and Julia Bradley from the University of Bristol in collaboration with Diabetes UK. This work is supported by the Elizabeth Blackwell Institute for Health Research, University of Bristol.

**Who has reviewed the study**

This study has been reviewed by the Faculty of Health Sciences Research Ethics Committee at the University of Bristol. For ethical enquiries about this study please contact .

**Further information and contact details**

**Online Survey**

Please note that respondents are only presented answers relevant to them, not the entire survey.

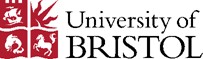

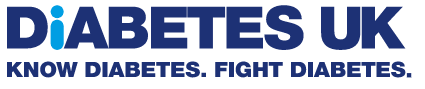

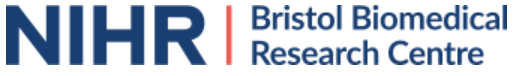
**Identifying support needs among people with diabetes during the Coronavirus COVID-19 pandemic**

Responsible researcher details: Dr Sarah Sauchelli Toran, NIHR Bristol Biomedical Research Centre-Nutrition, University of Bristol,

The NIHR Bristol Biomedical Research Centre is working with Diabetes UK to better understand and provide the type of support people with diabetes need during the coronavirus COVID-19 pandemic. Please read the information below before taking part.

**Who can take part?**

Adults aged 18 years or over who have a diagnosis of diabetes. Parents, carers or partners of people with diabetes can also take part.

**What will I be asked to do?**

You will be asked to complete a survey with questions about your well-being, the sources you use to obtain information/advice/support, your opinions on the advice you are receiving, and any improvements you would like to see in relation to guidance/advice.

This survey is completely anonymous. You are free to withdraw at any point. Any responses you have provided up to that point will be kept. This is because once entered, responses are automatically stored. As the survey is anonymous, we will not be able to extract the responses you have already provided. As sponsor for this study, the University of Bristol will manage your data securely in compliance with the General Data Protection Regulation for health and care research, and in accordance to the Data Protection Act 1998.

The information collected from this survey will be used to help Diabetes UK and may be used to support other research in the future. As part of a collaborative research approach to tackle the effects of coronavirus COVID-19, the data may be shared anonymously with other researchers.

If you wish to learn more about how your data will be stored and shared, please [**click here**](https://ords.onlinesurveys.ac.uk/diabetes-support-during-coronavirus-participant-informat).

- **I confirm that I am 18 years of age or older**
- **I confirm I have diabetes and / or am the parent/carer/partner of someone with diabetes**
- **I have read and understood the information on this page and give my consent to complete this survey**

If you do not wish to complete the survey, please exit this page.

If you have any questions or concerns about the survey, please contact us by emailing . If you would like to make a complaint about this survey, please contact **.**

**Introduction**

**We will distribute this survey again in the future to see if people’s experiences change over time. Please write in the box below a unique identifier that does not reveal your name (e.g. LEAF123) and remember it for the future. You can make a note of it. We will not be able to identify you through this but will be able to see changes across time.**

**This is optional and you can continue without entering an ID.**

[free text]

**Section A – Demographic characteristics**

First, we would like to know a few things about you.

| **Questions** | **Notes** |
| --- | --- |
| **What is your connection to diabetes?**  [ ] I have type 1 diabetes  [ ] I have type 2 diabetes  [ ] I have another type of diabetes  [ ] I am a parent or carer of someone with diabetes  [ ] I am the partner of someone with diabetes  [ ] I prefer not to say | ACTION: when 4^th^ or 5^th^ option selected, participants are directed to version of questions for parents/carers/partners |
| **Which part of the UK do you live in? Write down the first part of your postcode, the part before the space (e.g. SW14 for SW14 7QX).**  [ ] [ ] [ ] [ ] |  |
| **How old are you?**  [ ] [ ] [ ] | Number 18 to 112 |
| **What is your gender?**  [ ] Male  [ ] Female  [ ] Other |  |
| **What is your ethnicity?**  [ ] White: British  [ ] White: Irish  [ ] White: Gypsy or Irish Traveller  [ ] Other White background  [ ] Mixed: White and Black Caribbean  [ ] Mixed: White and Black African  [ ] Mixed: White and Asian  [ ] Other Mixed background  [ ] Asian or Asian British: Indian  [ ] Asian or Asian British: Pakistani  [ ] Asian or Asian British: Bangladeshi  [ ] Asian or Asian British: Chinese  [ ] Other Asian background  [ ] Black or Black British: African  [ ] Black or Black British: Caribbean  [ ] Other Black background  [ ] Arab  [ ] Other ethnic group  [ ] Prefer not to answer | From ONS. |

This page asks you a few more demographic questions related to Coronavirus

| **Questions** | **Notes** |
| --- | --- |
| **Are you currently living alone?**  [ ] Yes  [ ] No |  |
| **If No:**  **How many other adults (over 18 years) are you living with?** [ ] [ ]  **How many children (under 18 years) are you living with?** [ ] [ ] |  |
| **Has the number of people you are living with changed as a result of the coronavirus pandemic?**  [ ] Yes  [ ] No | From symptom tracking surveys being released. |
| **Which of the following best describes your current circumstances?**  [ ] I am following stringent social/physical distancing (e.g. reducing social contact but leaving the house for shopping and other essentials).  [ ] I am self-isolating at home, either because I have symptoms of coronavirus or someone in my household does. I do not leave the house.  [ ] I am self-isolating at home to protect someone in my household who is shielding. I do not leave the house.  [ ] I am in the shielding group who are being told to stay at home at all times and avoid contact (e.g. not leaving the home even for shopping).  [ ] I am shielding but I have not been identified as required to shield. I do not leave the house and avoid all contact.  [ ] I am a key worker/still leaving home to work.  [ ] Don’t know  [ ] Other. Please specify [free text] | From survey released by DUK for consistency. |
| **Have you been diagnosed with or displayed symptoms of coronavirus since the beginning of February?**  [ ] I have been diagnosed with coronavirus  [ ] I have shown symptoms  [ ] I have not shown symptoms  [ ] I am not sure |  |
| If selected I have shown symptoms:  **Please tick all of the symptoms that you have shown**   - Fever (temperature) - Cough - Shortness of breath - Headache - Runny nose/sneezing - Tiredness - Sore throat - Muscle aches - Diarrhoea - Vomiting - Loss of taste/smell |  |

**Section B: Diabetes management**

This section asks questions about your diabetes management before and after the pandemic.

**BEFORE** the coronavirus pandemic and social/physical distancing guidance I was confident that…

| **Questions** | **Notes** |
| --- | --- |
| **I was able to check my blood sugar if necessary. (0-10)**  Could not do at all Certain could do  [ ]0 [ ]1 [ ]2 [ ]3 [ ]4 [ ]5 [ ]6 [ ]7 [ ]8 [ ]9 [ ]10  [ ] Not applicable to me | Adapted from Confidence in Diabetes Self-Management Questionnaire following Patient and Public Involvement and Diabetes UK review |
| **I was able to correct my blood sugar when the sugar level was too high. (0-10)**  Could not do at all Certain could do  [ ]0 [ ]1 [ ]2 [ ]3 [ ]4 [ ]5 [ ]6 [ ]7 [ ]8 [ ]9 [ ]10  [ ] Not applicable to me |  |
| **I was able to correct my blood sugar when the sugar level was too low. (0-10)**  Could not do at all Certain could do  [ ]0 [ ]1 [ ]2 [ ]3 [ ]4 [ ]5 [ ]6 [ ]7 [ ]8 [ ]9 [ ]10  [ ] Not applicable to me |  |
| **I had a good understanding of my blood sugar levels and how to regulate these between HbA1c tests (if type 2 diabetes mellitus). (0-10)**  Could not do at all Certain could do  [ ]0 [ ]1 [ ]2 [ ]3 [ ]4 [ ]5 [ ]6 [ ]7 [ ]8 [ ]9 [ ]10  [] Not applicable to me |  |
| **I was able to choose the correct foods when necessary (e.g. when blood sugar level too low/high). (0-10)**  Could not do at all Certain could do  [ ]0 [ ]1 [ ]2 [ ]3 [ ]4 [ ]5 [ ]6 [ ]7 [ ]8 [ ]9 [ ]10  [ ] Not applicable to me |  |
| **I was able to keep my weight within a healthy range. (0-10)**  Could not do at all Certain could do  [ ]0 [ ]1 [ ]2 [ ]3 [ ]4 [ ]5 [ ]6 [ ]7 [ ]8 [ ]9 [ ]10  [ ] Not applicable to me |  |
| **I was able to examine my feet for cuts/ulcers or other changes. (0-10)**  Could not do at all Certain could do  [ ]0 [ ]1 [ ]2 [ ]3 [ ]4 [ ]5 [ ]6 [ ]7 [ ]8 [ ]9 [ ]10 |  |
| **I was able to follow a healthy eating pattern most of the time. (0-10)**  Could not do at all Certain could do  [ ]0 [ ]1 [ ]2 [ ]3 [ ]4 [ ]5 [ ]6 [ ]7 [ ]8 [ ]9 [ ]10 |  |
| **I was able to follow recommendations regarding physical activity. (0-10)**  Could not do at all Certain could do  [ ]0 [ ]1 [ ]2 [ ]3 [ ]4 [ ]5 [ ]6 [ ]7 [ ]8 [ ]9 [ ]10 |  |
| **I was able to take care of my mental wellbeing. (0-10)**  Could not do at all Certain could do  [ ]0 [ ]1 [ ]2 [ ]3 [ ]4 [ ]5 [ ]6 [ ]7 [ ]8 [ ]9 [ ]10 |  |

--- SEPARATION IN PRESENTATION--

**AT PRESENT**, I am confident that…

| **Questions** | **Notes** |
| --- | --- |
| **I am able to check my blood sugar if necessary. (0-10)**  Cannot do at all Certain can do  [ ]0 [ ]1 [ ]2 [ ]3 [ ]4 [ ]5 [ ]6 [ ]7 [ ]8 [ ]9 [ ]10  [ ] Not applicable to me | Adapted from Confidence in Diabetes Self-Management Questionnaire following Patient and Public Involvement and Diabetes UK review |
| **I am able to correct my blood sugar when the sugar level is too high. (0-10)**  Cannot do at all Certain can do  [ ]0 [ ]1 [ ]2 [ ]3 [ ]4 [ ]5 [ ]6 [ ]7 [ ]8 [ ]9 [ ]10  [ ] Not applicable to me |  |
| **I am able to correct my blood sugar when the sugar level is too low. (0-10)**  Cannot do at all Certain can do  [ ]0 [ ]1 [ ]2 [ ]3 [ ]4 [ ]5 [ ]6 [ ]7 [ ]8 [ ]9 [ ]10  [ ] Not applicable to me |  |
| **I have a good understanding of my blood sugar levels and how to regulate these between HbA1c tests (if type 2 diabetes mellitus). (0-10)**  Cannot do at all Certain can do  [ ]0 [ ]1 [ ]2 [ ]3 [ ]4 [ ]5 [ ]6 [ ]7 [ ]8 [ ]9 [ ]10  [ ] Not applicable to me |  |
| **I am able to choose the correct foods when necessary (e.g. when blood sugar level too low/high). (0-10)**  Cannot do at all Certain can do  [ ]0 [ ]1 [ ]2 [ ]3 [ ]4 [ ]5 [ ]6 [ ]7 [ ]8 [ ]9 [ ]10  [ ] Not applicable to me |  |
| **I am able to keep my weight within a healthy range. (0-10)**  Cannot do at all Certain can do  [ ]0 [ ]1 [ ]2 [ ]3 [ ]4 [ ]5 [ ]6 [ ]7 [ ]8 [ ]9 [ ]10  [ ] Not applicable to me |  |
| **I am able to examine my feet for cuts/ulcers or other changes. (0-10)**  Cannot do at all Certain can do  [ ]0 [ ]1 [ ]2 [ ]3 [ ]4 [ ]5 [ ]6 [ ]7 [ ]8 [ ]9 [ ]10 |  |
| **I am able to follow a healthy eating pattern most of the time. (0-10)**  Cannot do at all Certain can do  [ ]0 [ ]1 [ ]2 [ ]3 [ ]4 [ ]5 [ ]6 [ ]7 [ ]8 [ ]9 [ ]10 |  |
| **I am able to follow recommendations regarding physical activity. (0-10)**  Cannot do at all Certain can do  [ ]0 [ ]1 [ ]2 [ ]3 [ ]4 [ ]5 [ ]6 [ ]7 [ ]8 [ ]9 [ ]10 |  |
| **I am able to take care of my mental wellbeing. (0-10)**  Cannot do at all Certain can do  [ ]0 [ ]1 [ ]2 [ ]3 [ ]4 [ ]5 [ ]6 [ ]7 [ ]8 [ ]9 [ ]10 |  |

| **Questions** | **Notes** |
| --- | --- |
| **If you think your diabetes self-management has changed since the start of the pandemic, what do you think would help you get back on track?**  [Free text…] | No limit on word input. |
| **Have you had to cancel diabetes appointments and check-ups because of the pandemic?**  [ ] Yes  [ ] No |  |
| **If yes:**  **What impact has this had on your confidence and ability to self-manage?**  [Free text…] |  |

**Section C – Check-up**

The questions below refer to how you are feeling as you complete this survey. If you answer “yes” to any of the questions, please consider whether you would like to continue with the survey. If you are experiencing stress, you can contact the Diabetes UK helpline by phone 0345 123 2399 or e-mail. Contact details for Diabetes UK Scotland are: 0141 212 8710 or e-mail. You can also visit their online forum <https://www.diabetes.org.uk/how_we_help/community/diabetes-support-forum>.

| **Questions** | **Notes** |
| --- | --- |
| **Has completion of this survey increased your levels of stress/anxiety/worry?**  [ ] Yes  [ ] No | Adapted from previous NHS distress protocols. Original version softened as these questions were the ones generating stress to the PPI group |
| **Has completion of this survey made you feel like crying?**  [ ] Yes  [ ] No |  |
| **Has completion of this survey made you fearful?**  [ ] Yes  [ ] No |  |
| **Right now, are you shaking?**  [ ] Yes  [ ] No |  |

**Section D – Sources used for information/advice/support during the pandemic.**

In this section we want to find out what resources you have been using for advice/guidance/support **during the pandemic**.

| **Questions** | **Notes** |
| --- | --- |
| **Which of these resources have you used for guidance on how you should behave regarding social/physical distancing measures? (Tick all that apply)**  [ ] News channels (e.g. newspaper, radio, TV, website)  [ ] Public Health England gov.uk website  [ ] Diabetes UK website  [ ] NHS website  [ ] Other website  [ ] Twitter  [ ] Facebook  [ ] GP, diabetes specialist nurse or other healthcare professional  [ ] Family  [ ] Friends  [ ] Employer  [ ] Diabetes support group/network (via telephone, WhatsApp, text messages)  [ ] Other. Please specify: [free text] |  |
| If selected News channels:  **Which news channel(s)?**  [free text]  If selected Diabetes UK website:  **Which part(s) of the Diabetes UK website?**   - Coronavirus webpage - Online forum - Other. Please specify [free text]   If selected Other website:  **Which other website(s)?**  [free text]  If selected Twitter:  **Which part(s) of Twitter?**   - Diabetes UK page - Other. Please specify: [free text]   If selected Facebook:  **Which part(s) of Facebook?**   - Diabetes UK page - Diabetes support group - Other. Please specify: [free text] |  |
| **Which one have you used the most? (Tick one)**  [ ] News channels (e.g. newspaper, radio, TV, website)  [ ] Public Health England gov.uk website  [ ] Diabetes UK website  [ ] NHS website  [ ] Other website  [ ] Twitter  [ ] Facebook  [ ] GP, diabetes specialist nurse or other healthcare professional  [ ] Family  [ ] Friends  [ ] Employer  [ ] Diabetes support group/network (via telephone, WhatsApp, text messages)  [ ] Other. Please specify: [free text] |  |
| If selected News channels:  **Which news channel?**  [free text]  If selected Diabetes UK website:  **Which part of the Diabetes UK website?**   - Coronavirus webpage - Online forum - Other. Please specify [free text]   If selected Other website:  **Which other website?**  [free text]  If selected Twitter:  **Which part of Twitter?**   - Diabetes UK page - Other. Please specify: [free text]   If selected Facebook:  **Which part of Facebook?**   - Diabetes UK page - Diabetes support group   Other. Please specify: [free text] |  |
| **Which of these resources have you been using for guidance on general diabetes self-management since the start of the pandemic? (Tick all that apply)**  [ ] News channels (e.g. newspaper, radio, TV, website)  [ ] Public Health England gov.uk website  [ ] Diabetes UK website  [ ] NHS website  [ ] Other website  [ ] Twitter  [ ] Facebook  [ ] GP, diabetes specialist nurse or other healthcare professional  [ ] Family  [ ] Friends  [ ] Employer  [ ] Diabetes support group/network (via telephone, WhatsApp, text messages)  [ ] Other. Please specify: [free text] |  |
| If selected News channels:  **Which news channel(s)?**  [free text]  If selected Diabetes UK website:  **Which part(s) of the Diabetes UK website?**   - Coronavirus webpage - Online forum - Other. Please specify [free text]   If selected Other website:  **Which other website(s)?**  [free text]  If selected Twitter:  **Which part(s) of Twitter?**   - Diabetes UK page - Other. Please specify: [free text]   If selected Facebook:  **Which part(s) of Facebook?**   - Diabetes UK page - Diabetes support group   Other. Please specify: [free text] |  |
| **Which of these resources do you use to obtain emotional support? (Tick all that apply)**  [ ] Diabetes UK website – online forum  [ ] Diabetes UK Helpline  [ ] Facebook groups  [ ] GP, diabetes specialist nurse or other healthcare professional  [ ] Family  [ ] Friends  [ ] Neighbour  [ ] Employer  [ ] Diabetes support group/network (via telephone, WhatsApp, text messages)  [ ] Other. Please specify: [free text] |  |
| If selected Facebook groups:  **Which Facebook group(s)?**  [free text]  If selected GP, diabetes specialist nurse or other healthcare professional:  **Which healthcare professional(s)?**  [free text] |  |
| **If you are living alone, are you receiving support from people outside your household?**  [ ] Yes  [ ] No  [ ] Not applicable |  |
| **If yes:**  **Who are you receiving support from?**   - Family - Friends - Neighbours - Other. Please specify: [free text] |  |
| **How have the resources you use for guidance/support regarding diabetes self-management changed since the start of the pandemic?**  [free text] |  |
| **Which means do you use to obtain advice/guidance/support from outside your household? (tick all that apply)**  [ ] Not applicable  [ ] Telephone  [ ] Computer/laptop  [ ] Mobile phone (smartphone)  [ ] Someone in my household tells me about it  [ ] Other. Please specify [free text] |  |

**Section E – Opinions on information/advice/support received**

In this section we want your feedback on the information/advice/support you have received regarding diabetes management and social/physical distancing guidelines during the pandemic.

| **Questions** | **Notes** |
| --- | --- |
| **In general, how difficult or easy has it been for you to obtain INFORMATION/ADVICE applicable to you on the following?**  [ ] Very difficult [ ] Difficult [ ] Moderate [ ] Easy [ ] Very easy [ ] Not applicable to me.   - Glucose control - Diet - Physical activity - Medication - Emotional well-being - Diabetes management if showing symptoms of coronavirus - Social/physical distancing actions to take |  |
| **In general, how difficult or easy has it been for you to obtain SUPPORT applicable to you on the following?**  [ ] Very difficult [ ] Difficult [ ] Moderate [ ] Easy [ ] Very easy [ ] Not applicable to me.   - Glucose control - Diet - Physical activity - Medication - Emotional well-being - Diabetes management if showing symptoms of coronavirus - Social/physical distancing actions to take |  |
| **How would you rate the QUALITY of the information/advice/support from the following sources or channels?**  [ ] Very Poor [ ] Poor [ ] Average [ ] Good [ ] Very good [ ] Not applicable to me.   - Government (e.g. webpage/daily briefs) - Diabetes UK - Social Media - News channels (e.g. newspapers/TV news) - Friends - Family - Employer - Healthcare team |  |
| **If you have rated any of the above as very poor, poor or average, what improvements do you think should be made?**  **Please describe the improvements that you think should be made (type 'NA' if not applicable)**  [free text]   - Government (e.g. webpage/daily briefs) - Diabetes UK - Social Media - News channels (e.g. newspapers/TV news) - Friends - Family - Employer - Healthcare team |  |

For the next questions, please consider your current network of family, friends, contacts.

| **Questions** | **Notes** |
| --- | --- |
| **How would you rate their understanding of your CURRENT diabetes self-management needs?**  [ ] Very Poor  [ ] Poor  [ ] Average  [ ] Good  [ ] Very good  [ ] Not applicable to me |  |
| **How would you rate their support in your diabetes self-management during the pandemic?**  [ ] Very Poor  [ ] Poor  [ ] Average  [ ] Good  [ ] Very good  [ ] Not applicable to me |  |
| **How has the support you are receiving changed since before the pandemic? (0 = stayed the same)**  Decreased Increased  [ ]-5 [ ]-4 [ ]-3 [ ]-2 [ ]-1 [ ]0 [ ]1 [ ]2 [ ]3 [ ]4 [ ]5 |  |
| **In what ways do they CURRENTLY support your diabetes self-management? (Tick all that apply)**  [ ] Food shopping and/or preparation  [ ] Picking up medication  [ ] Essential travel  [ ] Monitoring blood glucose  [ ] Emotional support  [ ] Access to online resources (e.g. website, video meetings)  [ ] Prompting self-management behaviours (physical activity, foot checking etc.)  [ ] Other. Please specify [free text]  [ ] Not applicable to me |  |
| **If you are living with others, please rate how much of the support you are receiving comes from the people in your household. (0-10)**  None at all All  [ ] 0 [ ]1 [ ]2 [ ]3 [ ]4 [ ]5 [ ]6 [ ]7 [ ]8 [ ]9 [ ]10  [ ] Not applicable to me |  |

**Submit responses**

Please press 'Finish' to submit your responses to this survey.

**Final page**

Thank you for your completing the study. If you want support with issues related to any of the content in this survey, please contact the Diabetes UK helpline, visit their website or their online forum:

Diabetes UK main webpage: <https://www.diabetes.org.uk/>

Diabetes UK online forum: <https://www.diabetes.org.uk/how_we_help/community/diabetes-support-forum>

Diabetes UK helplines:

Diabetes UK coronavirus guidance: <https://www.diabetes.org.uk/about_us/news/coronavirus>

For government measures: [www.gov.uk/coronavirus](http://www.gov.uk/coronavirus)

If you have any questions or concerns about the survey, please contact us by emailing . If you would like to make a complaint about this survey, please contact **.**

__________________________________________________//_________________________________________________

**VERSION FOR CARER/PARENT/PARTNER**

Participants are led to this section if they have indicated that they are a parent, carer or partner of someone with diabetes.

**Section A**

| **Questions** | **Notes** |
| --- | --- |
| **Which part of the UK do you live in? Write down the first part of your postcode, the part before the space (e.g. SW14 for SW14 7QX)**  [ ] [ ] [ ] [ ] |  |
| **How old are you?**  [ ] [ ] [ ] | Number 18 to 112 |
| **What is your gender?**  [ ] Male  [ ] Female  [ ] Other |  |
| **What is your ethnicity?**  [ ] White: British  [ ] White: Irish  [ ] White: Gypsy or Irish Traveller  [ ] Other White background  [ ] Mixed: White and Black Caribbean  [ ] Mixed: White and Black African  [ ] Mixed: White and Asian  [ ] Other Mixed background  [ ] Asian or Asian British: Indian  [ ] Asian or Asian British: Pakistani  [ ] Asian or Asian British: Bangladeshi  [ ] Asian or Asian British: Chinese  [ ] Other Asian background  [ ] Black or Black British: African  [ ] Black or Black British: Caribbean  [ ] Other Black background  [ ] Arab  [ ] Other ethnic group  [ ] Prefer not to answer | Adapted from the ONS. |

This page asks you a few more demographic questions related to Coronavirus

| **Questions** | **Notes** |
| --- | --- |
| **Are you currently living with the person who has diabetes?**  [ ] Yes  [ ] No |  |
| **If yes,**  **How many other adults (over 18 years) are you living with?** [ ] [ ]  **How many children (under 18 years) are you living with?** [ ] [ ] |  |
| **Has the number of people you are living with changed as a result of the coronavirus pandemic?**  [ ] Yes  [ ] No | From symptom tracking surveys being released. |
| **Which of the following best describes your current circumstances?**  [ ] I am following stringent social/physical distancing (e.g. reducing social contact but leaving the house for shopping and other essentials).  [ ] I am self-isolating at home, either because I have symptoms of coronavirus or someone in my household does. I do not leave the house.  [ ] I am self-isolating at home to protect someone in my household who is shielding. I do not leave the house.  [ ] I am in the shielding group who are being told to stay at home at all times and avoid contact (e.g. not leaving the home even for shopping).  [ ] I am shielding but I have not been identified as required to shield. I do not leave the house and avoid all contact.  [ ] I am a key worker/still leaving home to work.  [ ] Don’t know  [ ] Other. Please specify [free text] |  |
| **Have you been diagnosed with or displayed symptoms of coronavirus since the beginning of February?**  [ ] I have been diagnosed with coronavirus  [ ] I have shown symptoms  [ ] I have not shown symptoms  [ ] I am not sure |  |
| If selected I have shown symptoms:  **Please tick all of the symptoms that you have shown**   - Fever (temperature) - Cough - Shortness of breath - Headache - Runny nose/sneezing - Tiredness - Sore throat - Muscle aches - Diarrhoea - Vomiting - Loss of taste/smell |  |
| **Has the person with diabetes been diagnosed with or displayed symptoms of coronavirus since the beginning of February?**  [ ] They have been diagnosed with coronavirus  [ ] They have shown symptoms  [ ] They have not shown symptoms  [ ] I am not sure |  |
| If selected They have shown symptoms:  **Please tick all of the symptoms that they have shown**   - Fever (temperature) - Cough - Shortness of breath - Headache - Runny nose/sneezing - Tiredness - Sore throat - Muscle aches - Diarrhoea - Vomiting - Loss of taste/smell |  |

**Section B: Diabetes management**

This section asks questions about your ability to support the person with diabetes in their management of their condition before and after the pandemic.

**BEFORE** the coronavirus pandemic and social/physical distancing guidance I was confident that…

| **Questions** | **Notes** |
| --- | --- |
| **I was able to help them check their blood sugar if necessary. (0-10)**  Could not do at all Certain could do  [ ]0 [ ]1 [ ]2 [ ]3 [ ]4 [ ]5 [ ]6 [ ]7 [ ]8 [ ]9 [ ]10  [ ] Not applicable | Adapted from previous NHS distress protocols. Original version softened as these questions were the ones generating stress to the PPI group |
| **I was able to help them correct their blood sugar when the sugar level was too high. (0-10)**  Could not do at all Certain could do  [ ]0 [ ]1 [ ]2 [ ]3 [ ]4 [ ]5 [ ]6 [ ]7 [ ]8 [ ]9 [ ]10  [ ] Not applicable |  |
| **I was able to help them correct their blood sugar when the sugar level was too low. (0-10)**  Could not do at all Certain could do  [ ]0 [ ]1 [ ]2 [ ]3 [ ]4 [ ]5 [ ]6 [ ]7 [ ]8 [ ]9 [ ]10  [ ] Not applicable |  |
| **I had a good understanding of blood sugar levels and how to help them regulate these between HbA1c tests (if type 2 diabetes mellitus). (0-10)**  Could not do at all Certain could do  [ ]0 [ ]1 [ ]2 [ ]3 [ ]4 [ ]5 [ ]6 [ ]7 [ ]8 [ ]9 [ ]10  [] Not applicable |  |
| **I was able to help them choose the correct foods when necessary (e.g. when blood sugar level too low/high). (0-10)**  Could not do at all Certain could do  [ ]0 [ ]1 [ ]2 [ ]3 [ ]4 [ ]5 [ ]6 [ ]7 [ ]8 [ ]9 [ ]10  [ ] Not applicable |  |
| **I was able to help them keep their weight within a healthy range. (0-10)**  Could not do at all Certain could do  [ ]0 [ ]1 [ ]2 [ ]3 [ ]4 [ ]5 [ ]6 [ ]7 [ ]8 [ ]9 [ ]10  [ ] Not applicable |  |
| **I was able to help them examine their feet for cuts/ulcers or other changes. (0-10)**  Could not do at all Certain could do  [ ]0 [ ]1 [ ]2 [ ]3 [ ]4 [ ]5 [ ]6 [ ]7 [ ]8 [ ]9 [ ]10 |  |
| **I was able to help them follow a healthy eating pattern most of the time. (0-10)**  Could not do at all Certain could do  [ ]0 [ ]1 [ ]2 [ ]3 [ ]4 [ ]5 [ ]6 [ ]7 [ ]8 [ ]9 [ ]10 |  |
| **I was able to help them follow recommendations regarding physical activity. (0-10)**  Could not do at all Certain could do  [ ]0 [ ]1 [ ]2 [ ]3 [ ]4 [ ]5 [ ]6 [ ]7 [ ]8 [ ]9 [ ]10 |  |
| **I was able to help them take care of their mental wellbeing. (0-10)**  Could not do at all Certain could do  [ ]0 [ ]1 [ ]2 [ ]3 [ ]4 [ ]5 [ ]6 [ ]7 [ ]8 [ ]9 [ ]10 |  |

--- SEPARATION IN PRESENTATION--

**AT PRESENT**, I am confident that…

| **Questions** | **Notes** |
| --- | --- |
| **I am able to help them check their blood sugar if necessary. (0-10)**  Cannot do at all Certain can do  [ ]0 [ ]1 [ ]2 [ ]3 [ ]4 [ ]5 [ ]6 [ ]7 [ ]8 [ ]9 [ ]10  [ ] Not applicable | Adapted from previous NHS distress protocols. Original version softened as these questions were the ones generating stress to the PPI group |
| **I am able to help them correct their blood sugar when the sugar level is too high. (0-10)**  Cannot do at all Certain can do  [ ]0 [ ]1 [ ]2 [ ]3 [ ]4 [ ]5 [ ]6 [ ]7 [ ]8 [ ]9 [ ]10  [ ] Not applicable |  |
| **I am able to help them correct their blood sugar when the sugar level is too low. (0-10)**  Cannot do at all Certain can do  [ ]0 [ ]1 [ ]2 [ ]3 [ ]4 [ ]5 [ ]6 [ ]7 [ ]8 [ ]9 [ ]10  [ ] Not applicable |  |
| **I have a good understanding of blood sugar levels and how to help them regulate these between HbA1c tests (if type 2 diabetes mellitus). (0-10)**  Cannot do at all Certain can do  [ ]0 [ ]1 [ ]2 [ ]3 [ ]4 [ ]5 [ ]6 [ ]7 [ ]8 [ ]9 [ ]10  [] Not applicable |  |
| **I am able to help them choose the correct foods when necessary (e.g. when blood sugar level too low/high). (0-10)**  Cannot do at all Certain can do  [ ]0 [ ]1 [ ]2 [ ]3 [ ]4 [ ]5 [ ]6 [ ]7 [ ]8 [ ]9 [ ]10  [ ] Not applicable |  |
| **I am able to help them keep their weight within a healthy range. (0-10)**  Cannot do at all Certain can do  [ ]0 [ ]1 [ ]2 [ ]3 [ ]4 [ ]5 [ ]6 [ ]7 [ ]8 [ ]9 [ ]10 |  |
| **I am able to help them examine their feet for cuts/ulcers or other changes. (0-10)**  Cannot do at all Certain can do  [ ]0 [ ]1 [ ]2 [ ]3 [ ]4 [ ]5 [ ]6 [ ]7 [ ]8 [ ]9 [ ]10 |  |
| **I am able to help them follow a healthy eating pattern most of the time. (0-10)**  Cannot do at all Certain can do  [ ]0 [ ]1 [ ]2 [ ]3 [ ]4 [ ]5 [ ]6 [ ]7 [ ]8 [ ]9 [ ]10 |  |
| **I am able to help them follow recommendations regarding physical activity. (0-10)**  Cannot do at all Certain can do  [ ]0 [ ]1 [ ]2 [ ]3 [ ]4 [ ]5 [ ]6 [ ]7 [ ]8 [ ]9 [ ]10 |  |
| **I am able to help them take care of their mental wellbeing. (0-10)**  Cannot do at all Certain can do  [ ]0 [ ]1 [ ]2 [ ]3 [ ]4 [ ]5 [ ]6 [ ]7 [ ]8 [ ]9 [ ]10 |  |

| **Questions** | **Notes** |
| --- | --- |
| **If you think your ability to provide support in diabetes self-management has changed since the start of the pandemic, what do you think would help you improve the support you can provide?**  [Free text…] | No limit on word input. |

**Section C – Check-up**

The questions below refer to how you are feeling as you complete this survey. If you answer “yes” to any of the questions, please consider whether you would like to continue with the survey. If you are experiencing stress, you can contact the Diabetes UK helpline by phone 0345 123 2399 or e-mail. Contact details for Diabetes UK Scotland are: 0141 212 8710 or e-mail. You can also visit their online forum <https://www.diabetes.org.uk/how_we_help/community/diabetes-support-forum>.

| **Questions** | **Notes** |
| --- | --- |
| **Has completion of this survey increased your levels of stress/anxiety/worry?**  [ ] Yes  [ ] No | Adapted from previous NHS distress protocols. Original version softened as these questions were the ones generating stress to the PPI group. |
| **Has completion of this survey made you feel like crying?**  [ ] Yes  [ ] No |  |
| **Has completion of this survey made you fearful?**  [ ] Yes  [ ] No |  |
| **Right now, are you shaking?**  [ ] Yes  [ ] No |  |

**Section D – Sources used for information/advice/support during the pandemic.**

In this section we want to find out what resources you have been using for advice/guidance/support **during the pandemic**.

| **Questions** | **Notes** |
| --- | --- |
| **Which of these resources have you used for guidance on how you should behave regarding social/physical distancing measures in relation to the person with diabetes? (Tick all that apply)**  [ ] News channels (e.g. newspaper, radio, TV, website)  [ ] Public Health England gov.uk website  [ ] Diabetes UK website  [ ] NHS website  [ ] Other website  [ ] Twitter  [ ] Facebook  [ ] GP, diabetes specialist nurse or other healthcare professional  [ ] Family  [ ] Friends  [ ] Employer  [ ] Diabetes support group/network (via telephone, WhatsApp, text messages)  [ ] Other. Please specify: [free text] |  |
| If selected News channels:  **Which news channel(s)?**  [free text]  If selected Diabetes UK website:  **Which part(s) of the Diabetes UK website?**   - Coronavirus webpage - Online forum - Other. Please specify [free text]   If selected Other website:  **Which other website(s)?**  [free text]  If selected Twitter:  **Which part(s) of Twitter?**   - Diabetes UK page - Other. Please specify: [free text]   If selected Facebook:  **Which part(s) of Facebook?**   - Diabetes UK page - Diabetes support group - Other. Please specify: [free text] |  |
| **Which one have you use the most? (Tick one)**  [ ] News channels (e.g. newspaper, radio, TV, website)  [ ] Public Health England gov.uk website  [ ] Diabetes UK website  [ ] NHS website  [ ] Other website  [ ] Twitter  [ ] Facebook  [ ] GP, diabetes specialist nurse or other healthcare professional  [ ] Family  [ ] Friends  [ ] Employer  [ ] Diabetes support group/network (via telephone, WhatsApp, text messages)  [ ] Other. Please specify: [free text] |  |
| If selected News channels:  **Which news channel?**  [free text]  If selected Diabetes UK website:  **Which part of the Diabetes UK website?**   - Coronavirus webpage - Online forum - Other. Please specify [free text]   If selected Other website:  **Which other website?**  [free text]  If selected Twitter:  **Which part of Twitter?**   - Diabetes UK page - Other. Please specify: [free text]   If selected Facebook:  **Which part of Facebook?**   - Diabetes UK page - Diabetes support group - Other. Please specify: [free text] |  |
| **Which of these resources have you been using for guidance on general diabetes management since the start of the pandemic? (Tick all that apply)**  [ ] News channels (e.g. newspaper, radio, TV, website)  [ ] Public Health England gov.uk website  [ ] Diabetes UK website  [ ] NHS website  [ ] Other website  [ ] Twitter  [ ] Facebook  [ ] GP, diabetes specialist nurse or other healthcare professional  [ ] Family  [ ] Friends  [ ] Employer  [ ] Diabetes support group/network (via telephone, WhatsApp, text messages)  [ ] Other. Please specify: [free text] |  |
| If selected News channels:  **Which news channel(s)?**  [free text]  If selected Diabetes UK website:  **Which part(s) of the Diabetes UK website?**   - Coronavirus webpage - Online forum - Other. Please specify [free text]   If selected Other website:  **Which other website(s)?**  [free text]  If selected Twitter:  **Which part(s) of Twitter?**   - Diabetes UK page - Other. Please specify: [free text]   If selected Facebook:  **Which part(s) of Facebook?**   - Diabetes UK page - Diabetes support group - Other. Please specify: [free text] |  |
| **Which of these resources do you use to obtain emotional support? (Tick all that apply)**  [ ] Diabetes UK website – online forum  [ ] Diabetes UK Helpline  [ ] Facebook groups  [ ] GP, diabetes specialist nurse or other healthcare professional  [ ] Family  [ ] Friends  [ ] Employer  [ ] Diabetes support group/network (via telephone, WhatsApp, text messages)  [ ] Other. Please specify: [free text] |  |
| If selected Facebook groups:  **Which Facebook group(s)?**  [free text]  If selected GP, diabetes specialist nurse or other healthcare professional:  **Which healthcare professional(s)?**  [free text] |  |
| **How have the resources you use for guidance/support on how to help in diabetes management changed since the start of the pandemic?**  [free text] |  |
| **Which means do you use to obtain advice/guidance/support from outside your household? (tick all that apply)**  [ ] Not applicable  [ ] Telephone  [ ] Computer/laptop  [ ] Mobile phone (smartphone)  [ ] Someone in my household tells me about it  [ ] Other. Please specify [free text] |  |

**Section E – Opinions on information/advice/support received**

In this section we want your feedback on the information/advice/support you have received regarding diabetes management and social/physical distancing guidelines during the pandemic.

| **Questions** | **Notes** |
| --- | --- |
| **In general, how difficult or easy has it been for you to obtain INFORMATION/ADVICE applicable to the person you are helping on the following?**  [ ] Very difficult [ ] Difficult [ ] Moderate [ ] Easy [ ] Very easy [ ] Not applicable to me.   - Glucose control - Diet - Physical activity - Medication - Emotional well-being - Diabetes management if showing symptoms of coronavirus - Social/physical distancing actions to take |  |
| **In general, how difficult or easy has it been for you to obtain SUPPORT applicable to the person you are helping on the following?**  [ ] Very difficult [ ] Difficult [ ] Moderate [ ] Easy [ ] Very easy [ ] Not applicable to me.   - Glucose control - Diet - Physical activity - Medication - Emotional well-being - Diabetes management if showing symptoms of coronavirus - Social/physical distancing actions to take |  |
| **How would you rate the QUALITY of the information/advice/support from the following sources or channels?**  [ ] Very Poor [ ] Poor [ ] Average [ ] Good [ ] Very good [ ] Not applicable to me.   - Government (e.g. webpage/daily briefs) - Diabetes UK - Social Media - News channels (e.g. newspapers/TV news) - Friends - Family - Employer - Healthcare team |  |
| **If you have rated any of the above as very poor, poor or average, what improvements do you think should be made?**  **Please describe the improvements that you think should be made (type 'NA' if not applicable)**  [free text]   - Government (e.g. webpage/daily briefs) - Diabetes UK - Social Media - News channels (e.g. newspapers/TV news) - Friends - Family - Employer - Healthcare team |  |

For the next questions, please reflect on your role as someone helping an individual with diabetes.

| **Questions** | **Notes** |
| --- | --- |
| **How would you rate your understanding of their CURRENT diabetes self-management needs?**  [ ] Very Poor  [ ] Poor  [ ] Average  [ ] Good  [ ] Very good  [ ] Not applicable to me |  |
| **In what ways do you CURRENTLY support the individual in their diabetes self-management? (Tick all that apply)**  [ ] Food shopping and/or preparation  [ ] Picking up medication  [ ] Essential travel  [ ] Monitoring blood glucose  [ ] Emotional support  [ ] Access to online resources (e.g. website, video meetings)  [ ] Prompting self-management behaviours (physical activity, foot checking etc.)  [ ] Other. Please specify [free text]  [ ] Not applicable to me | Feel free to add any other response. |

**Submit responses**

Please press 'Finish' to submit your responses to this survey.

**Final page**

Thank you for your completing the study. If you want support with issues related to any of the content in this survey, please contact the Diabetes UK helpline, visit their website or their online forum:

Diabetes UK main webpage: <https://www.diabetes.org.uk/>

Diabetes UK online forum: <https://www.diabetes.org.uk/how_we_help/community/diabetes-support-forum>

Diabetes UK helplines:

Diabetes UK coronavirus guidance: <https://www.diabetes.org.uk/about_us/news/coronavirus>

For government measures: [www.gov.uk/coronavirus](http://www.gov.uk/coronavirus)

If you have any questions or concerns about the survey, please contact us by emailing . If you would like to make a complaint about this survey, please contact 
