## Supplementary File 2 for "Exploring support needs of people living with diabetes during the coronavirus COVID-19 pandemic: insights from a UK survey"

**Responses to the survey**

### 1. **Responses from people living with diabetes**

#### 1.1. Geographical distribution of responses

| **Region/Nation** | **n** | **%** |
| --- | --- | --- |
| Scotland | 89 | 11.6 |
| Wales | 32 | 4.18 |
| East Midlands | 31 | 4.05 |
| East of England | 52 | 6.8 |
| Greater London | 65 | 8.5 |
| North East | 14 | 1.83 |
| North West | 55 | 7.19 |
| Northern Ireland | 14 | 1.83 |
| South East | 193 | 25.2 |
| South West | 142 | 18.6 |
| West Midlands | 57 | 7.45 |
| Yorkshire & Humber | 21 | 2.75 |
| **Total** | **765** |  |

#### 1.2. Demographic characteristics.

| **Diabetes group** | **n** | **%** |
| --- | --- | --- |
| Type 1 diabetes | 535 | 69.2 |
| Type 2 diabetes | 220 | 28.5 |
| Another type of diabetes | 18 | 2.3 |
|  | 773 | 100.0 |

|  | **All**  **(n=773)** | **Type 1**  **(n=535)** | **Type 2**  **(n=220)** |
| --- | --- | --- | --- |
| **Gender, n (%)** |  |  |  |
| Female | 516 (67.1%) | 365 (68.6%) | 139 (63.5%) |
| Male | 249 (32.4%) | 165 (31.0%) | 78 (35.6%) |
| Other | 4 (0.5%) | 2 (0.4%) | 2 (0.9%) |
| **Age, mean (SD)** | 47.9 (14.5) | 44.4 (14.2) | 56.5 (11.4) |
| **Ethnicity, n (%)** |  |  |  |
| Arab | 1 (0.1%) | 1 (0.2%) | 0 (0.0%) |
| Asian or Asian British: Chinese | 3 (0.4%) | 0 (0.0%) | 3 (1.4%) |
| Asian or Asian British: Indian | 8 (1.0%) | 2 (0.4%) | 6 (2.7%) |
| Asian or Asian British: Pakistani | 1 (0.1%) | 1 (0.2%) | 0 (0.0%) |
| Black or Black British: Caribbean | 4 (0.5%) | 0 (0.0%) | 4 (1.8%) |
| Mixed: White and Asian | 5 (0.7%) | 3 (0.6%) | 2 (0.9%) |
| Mixed: White and Black African | 1 (0.1%) | 1 (0.2%) | 0 (0.0%) |
| Mixed: White and Black Caribbean | 1 (0.1%) | 1 (0.2%) | 0 (0.0%) |
| Other ethnic group | 1 (0.1%) | 0 (0.0%) | 1 (0.5%) |
| Other Mixed background | 1 (0.1%) | 1 (0.2%) | 0 (0.0%) |
| Other White background | 31 (4.0%) | 26 (4.9%) | 5 (2.3%) |
| Prefer not to answer | 3 (0.4%) | 2 (0.4%) | 1 (0.5%) |
| White: British | 693 (90.1%) | 485 (91.2%) | 192 (87.7%) |
| White: Irish | 16 (2.1%) | 9 (1.7%) | 5 (2.3%) |

**Living circumstances**

| **Are you currently living alone?** | **All** | **Type 1** | **Type 2** |
| --- | --- | --- | --- |
| No | 649 (84.1%) | 458 (85.6%) | 176 (80.4%) |
| Yes | 123 (15.9%) | 77 (14.4%) | 43 (19.6%) |
|  | 772 (100.0%) | 535 (100.0%) | 219 (100.0%) |

| **Has the number of people you are living with changed as a result of the coronavirus pandemic?** | **All** | **Type 1** | **Type 2** |
| --- | --- | --- | --- |
| No | 684 (88.9%) | 467 (87.6%) | 200 (91.7%) |
| Yes | 85 (11.1%) | 66 (12.4%) | 18 (8.3%) |
|  | 769 (100.0%) | 533 (100.0%) | 218 (100.0%) |

**Circumstances in relation to COVID-19**

| **Have you been diagnosed with or displayed symptoms of coronavirus since the beginning of February?** | **All** | **Type 1** | **Type 2** |
| --- | --- | --- | --- |
| No | 623 (81.0%) | 434 (81.8%) | 176 (80.0%) |
| Yes | 70 (9.1%) | 47 (8.9%) | 21 (9.5%) |
| Diagnosed with coronavirus | 2 (0.3%) | 2 (0.4%) | 0 (0.0%) |
| Not sure | 74 (9.6%) | 48 (9.0%) | 23 (10.5%) |
|  | 769 (100.0%) | 531 (100.0%) | 220 (100.0%) |

| **Which of the following best describes your current circumstances?** | **All** | **Type 1** | **Type 2** |
| --- | --- | --- | --- |
| Following stringent Physical/social/physical distancing | 513 (66.8%) | 355 (66.9%) | 147 (67.1%) |
| Self-isolating at home | 16 (2.1%) | 9 (1.7%) | 7 (3.3%) |
| Shielding group | 59 (7.7%) | 37 (7.0%) | 19 (8.7%) |
| Shielding (but not in shielding group) | 75 (9.8%) | 49 (9.2%) | 22 (10.0%) |
| Key worker/still leaving home to work | 97 (12.6%) | 75 (14.1%) | 22 (10.1%) |
| Other | 4 (5.7%) | 3 (0.6%) | 1 (0.5%) |
| Don’t know | 4 (0.5%) | 3 (0.6%) | 1 (0.5%) |
|  | 768 (100.0%) | 531 (100.0%) | 219 (100.0%) |

#### 1.3. Confidence in diabetes self-management

| **BEFORE** the coronavirus pandemic and social/physical distancing guidance I was confident that I was able to… | **All** |  | **Type 1** |  | **Type 2** |  |
| --- | --- | --- | --- | --- | --- | --- |
|  | n | Median (IQR) | n | Median (IQR) | n | Median (IQR) |
| Check blood glucose | 720 | 10 (10, 10) | 528 | 10 (10, 10) | 174 | 10 (9, 10) |
| Correct high blood glucose | 703 | 10 (8, 10) | 530 | 10 (9, 10) | 156 | 7 (3, 10) |
| Correct low blood glucose | 680 | 10 (9, 10) | 526 | 10 (10, 10) | 136 | 10 (7, 10) |
| Good blood glucose regulation | 571 | 9 (7, 10) | 363 | 10 (8, 10) | 197 | 8 (5, 10) |
| Choose correct foods | 729 | 10 (8, 10) | 519 | 10 (9, 10) | 192 | 8 (5, 10) |
| Keep healthy weight | 755 | 7 (4, 10) | 525 | 8 (5, 10) | 212 | 5 (2, 8) |
| Examine feet | 769 | 10 (8, 10) | 533 | 10 (8, 10) | 218 | 10 (7, 10) |
| Healthy eating pattern | 770 | 8 (6, 10) | 534 | 9 (7, 10) | 218 | 7 (5, 9) |
| Physical activity | 770 | 8 (5, 10) | 534 | 8 (6, 10) | 218 | 6 (4, 9) |
| Mental wellbeing | 770 | 8 (5, 10) | 534 | 8 (6, 10) | 218 | 7 (5, 10) |

Note: score given on a Likert scale ranging from 0 (Could not do at all) to 10 (Certain could do). Not applicable was also an option to account inter-individual variability in condition and self-management requirements.

| **AT PRESENT**, I am confident that… | **All** |  | **Type 1** |  | **Type 2** |  |
| --- | --- | --- | --- | --- | --- | --- |
|  | n | Median (IQR) | N | Median (IQR) | n | Median (IQR) |
| Check blood glucose | 727 | 10 (10, 10) | 530 | 10 (10, 10) | 179 | 10 (9, 10) |
| Correct high blood glucose | 718 | 10 (8, 10) | 531 | 10 (9, 10) | 170 | 7 (4, 10) |
| Correct low blood glucose | 697 | 10 (9, 10) | 526 | 10 (9, 10) | 153 | 9 (6, 10) |
| Good blood glucose regulation | 578 | 9 (7, 10) | 367 | 10 (8, 10) | 200 | 8 (5, 10) |
| Choose correct foods | 744 | 9 (7, 10) | 524 | 10 (8, 10) | 202 | 8 (5, 10) |
| Keep healthy weight | 759 | 7 (4, 9) | 524 | 7 (5, 10) | 217 | 5 (2, 7) |
| Examine feet | 764 | 10 (8, 10) | 527 | 10 (8, 10) | 219 | 10 (7, 10) |
| Healthy eating pattern | 766 | 8 (6, 10) | 530 | 8 (6, 10) | 218 | 7 (5, 9) |
| Physical activity | 768 | 7 (5, 10) | 531 | 8 (5, 10) | 219 | 6 (4, 8) |
| Mental wellbeing | 765 | 7 (5, 9) | 530 | 7 (5, 9) | 217 | 7 (4, 9) |

Note: score given on a Likert scale ranging from 0 (Could not do at all) to 10 (Certain could do). Not applicable was also an option to account inter-individual variability in condition and self-management requirements.

| **Change in score** | **All** | **Type 1** | **Type 2** |
| --- | --- | --- | --- |
| **Check blood glucose** |  |  |  |
| Decreased | 68 (9.6%) | 45 (8.6%) | 23 (13.5%) |
| Same | 600 (84.3%) | 453 (86.5%) | 129 (75.9%) |
| Increased | 44 (6.2%) | 26 (5.0%) | 18 (10.6%) |
| **Correct high blood glucose** |  |  |  |
| Decreased | 95 (13.6%) | 67 (12.7%) | 25 (16.2%) |
| Same | 510 (73.1%) | 410 (77.8%) | 88 (57.1%) |
| Increased | 93 (13.3%) | 50 (9.5%) | 41 (26.6%) |
| **Correct low blood glucose** |  |  |  |
| Decreased | 92 (13.7%) | 71 (13.7%) | 20 (14.8%) |
| Same | 522 (77.7%) | 412 (79.4%) | 93 (68.9%) |
| Increased | 58 (8.6%) | 36 (6.9%) | 22 (16.3%) |
| **Good blood glucose regulation** |  |  |  |
| Decreased | 92 (16.6%) | 48 (13.5%) | 42 (22.1%) |
| Same | 380 (68.5%) | 262 (73.8%) | 111 (58.4%) |
| Increased | 83 (15.0%) | 45 (12.7%) | 37 (19.5%) |
| **Choose correct foods** |  |  |  |
| Decreased | 173 (24.1%) | 107 (20.8%) | 60 (32.1%) |
| Same | 440 (61.2%) | 345 (67.1%) | 87 (46.5%) |
| Increased | 106 (14.7%) | 62 (12.1%) | 40 (21.4%) |
| **Keep healthy weight** |  |  |  |
| Decreased | 202 (27.1%) | 144 (27.9%) | 53 (25.2%) |
| Same | 386 (51.8%) | 284 (54.9%) | 93 (44.3%) |
| Increased | 157 (21.1%) | 89 (17.2%) | 64 (30.5%) |
| **Examine feet** |  |  |  |
| Decreased | 87 (11.4%) | 58 (11.0%) | 27 (12.4%) |
| Same | 598 (78.7%) | 419 (79.8%) | 163 (75.1%) |
| Increased | 75 (9.9%) | 48 (9.1%) | 27 (12.4%) |
| **Healthy eating pattern** |  |  |  |
| Decreased | 226 (29.6%) | 153 (28.9%) | 69 (31.9%) |
| Same | 393 (51.5%) | 287 (54.3%) | 93 (43.1%) |
| Increased | 144 (18.9%) | 89 (16.8%) | 54 (25.0%) |
| **Physical activity** |  |  |  |
| Decreased | 245 (32.0%) | 166 (31.3%) | 71 (32.7%) |
| Same | 361 (47.2%) | 257 (48.5%) | 96 (44.2%) |
| Increased | 159 (20.8%) | 107 (20.2%) | 50 (23.0%) |
| **Mental wellbeing** |  |  |  |
| Decreased | 282 (37.0%) | 207 (39.1%) | 66 (30.7%) |
| Same | 362 (47.5%) | 244 (46.1%) | 110 (51.2%) |
| Increased | 118 (15.5%) | 78 (14.7%) | 39 (18.1%) |

**Difference between respondents living alone and those living with others**

| **Change in score** | **Not living alone** | **Living alone** |
| --- | --- | --- |
| **Check blood glucose** |  |  |
| Decreased | 55 (9.0%) | 13 (12.6%) |
| Same | 516 (84.9%) | 84 (81.6%) |
| Increased | 37 (6.1%) | 6 (5.8%) |
| **Correct high blood glucose** |  |  |
| Decreased | 71 (11.9%) | 24 (23.3%) |
| Same | 445 (74.8%) | 65 (63.1%) |
| Increased | 79 (13.3%) | 14 (13.6%) |
| **Correct low blood glucose** |  |  |
| Decreased | 70 (12.3%) | 22 (21.8%) |
| Same | 451 (79.0%) | 71 (70.3%) |
| Increased | 50 (8.8%) | 8 (7.9%) |
| **Good blood glucose regulation** |  |  |
| Decreased | 74 (15.6%) | 18 (22.0%) |
| Same | 327 (69.1%) | 53 (64.6%) |
| Increased | 72 (15.2%) | 11 (13.4%) |
| **Choose correct foods** |  |  |
| Decreased | 150 (24.6%) | 23 (20.9%) |
| Same | 373 (61.2%) | 67 (60.9%) |
| Increased | 86 (14.1%) | 20 (18.2%) |
| **Keep healthy weight** |  |  |
| Decreased | 166 (26.3%) | 36 (31.6%) |
| Same | 329 (52.2%) | 57 (50.0%) |
| Increased | 135 (21.4%) | 21 (18.4%) |
| **Examine feet** |  |  |
| Decreased | 68 (10.6%) | 19 (16.1%) |
| Same | 508 (79.3%) | 90 (76.3%) |
| Increased | 65 (10.1%) | 9 (7.6%) |
| **Healthy eating pattern** |  |  |
| Decreased | 185 (28.7%) | 41 (35.0%) |
| Same | 335 (51.9%) | 57 (48.7%) |
| Increased | 125 (19.4%) | 19 (16.2%) |
| **Physical activity** |  |  |
| Decreased | 190 (29.5%) | 55 (46.2%) |
| Same | 309 (47.9%) | 51 (42.9%) |
| Increased | 146 (22.6%) | 13 (10.9%) |
| **Mental wellbeing** |  |  |
| Decreased | 235 (36.6%) | 47 (39.5%) |
| Same | 301 (46.9%) | 60 (50.4%) |
| Increased | 106 (16.5%) | 12 (10.1%) |

**Differences between individuals living alone that were not receiving outside support and those that were.**

| **Change in score** | **Not receiving outside support** | **Receiving outside support** |
| --- | --- | --- |
| **Check blood glucose** |  |  |
| Decreased | 7 (17%) | 5 (9%) |
| Same | 30 (73%) | 51 (89%) |
| Increased | 4 (10%) | 1 (2%) |
| **Correct high blood glucose** |  |  |
| Decreased | 10 (25%) | 14 (24%) |
| Same | 23 (57%) | 39 (66%) |
| Increased | 7 (18%) | 6 (10%) |
| **Correct low blood glucose** |  |  |
| Decreased | 6 (16%) | 15 (26%) |
| Same | 30 (79%) | 38 (66%) |
| Increased | 2 (5%) | 5 (9%) |
| **Good blood glucose regulation** |  |  |
| Decreased | 7 (22%) | 10 (22%) |
| Same | 19 (59%) | 33 (72%) |
| Increased | 6 (19%) | 3 (7%) |
| **Choose correct foods** |  |  |
| Decreased | 8 (19%) | 15 (24%) |
| Same | 24 (56%) | 40 (63%) |
| Increased | 11 (26%) | 8 (13%) |
| **Keep healthy weight** |  |  |
| Decreased | 10 (23%) | 24 (37%) |
| Same | 21 (49%) | 33 (51%) |
| Increased | 12 (28%) | 8 (12%) |
| **Examine feet** |  |  |
| Decreased | 5 (11%) | 13 (19%) |
| Same | 33 (75%) | 52 (78%) |
| Increased | 6 (14%) | 2 (3%) |
| **Healthy eating pattern** |  |  |
| Decreased | 15 (34%) | 23 (35%) |
| Same | 21 (48%) | 32 (48%) |
| Increased | 8 (18%) | 11 (17%) |
| **Physical activity** |  |  |
| Decreased | 18 (40%) | 34 (51%) |
| Same | 23 (51%) | 24 (36%) |
| Increased | 4 (9%) | 9 (13%) |
| **Mental wellbeing** |  |  |
| Decreased | 18 (40%) | 27 (40%) |
| Same | 25 (56%) | 31 (46%) |
| Increased | 2 (4%) | 9 (13%) |

Note: This table is restricted to participants who reported living alone for the question on living circumstances.

Qualitative responses regarding what respondents perceive they need to improve their diabetes self-management are summarised in the main manuscript.

#### 1.4. Cancellation of clinical appointments

| **Have you had to cancel diabetes appointments and check-ups because of the pandemic?** | **All** | **Type 1** | **Type 2** |
| --- | --- | --- | --- |
| No | 372 (48.2%) | 249 (46.7%) | 118 (53.6%) |
| Yes | 399 (51.8%) | 284 (53.3%) | 102 (46.4%) |
|  | 771 (100.0%) | 533 (100.0%) | 220 (100.0%) |

Qualitative responses regarding the impact of the cancellation of appointments are summarised in the appendices.

#### 1.5. Sources used for information, advice and support

| **Which of these resources have you used for guidance on how you should behave regarding social/physical distancing measures? (Tick all that apply)** | **All**  **(n = 770)** | **%** | **Type 1**  **(n = 535)** | **%** | **Type 2**  **(n = 217)** | **%** |
| --- | --- | --- | --- | --- | --- | --- |
| News channels | 557 | 72.3 | 384 | 71.8 | 158 | 72.8 |
| Public Health and government website | 386 | 50.1 | 286 | 53.5 | 91 | 41.9 |
| Diabetes UK website | 461 | 59.9 | 340 | 63.6 | 110 | 50.7 |
| NHS website | 386 | 50.1 | 282 | 52.7 | 100 | 46.1 |
| Other website | 52 | 6.8 | 36 | 6.7 | 12 | 5.5 |
| Physical/social media | 219 | 28.4 | 156 | 29.2 | 56 | 25.8 |
| GP, diabetes nurse, healthcare professional | 261 | 33.9 | 185 | 34.6 | 68 | 31.3 |
| Family | 174 | 22.6 | 125 | 23.4 | 41 | 18.9 |
| Friends | 111 | 14.4 | 79 | 14.8 | 28 | 12.9 |
| Employer | 113 | 14.7 | 85 | 15.9 | 23 | 10.6 |
| Diabetes support group | 67 | 8.7 | 52 | 9.7 | 11 | 5.1 |
| Other | 12 | 1.6 | 7 | 1.3 | 4 | 1.8 |

| **Which one have you used the most? (Tick one)** | **All** | **%** | **Type 1** | **%** | **Type 2** | **%** |
| --- | --- | --- | --- | --- | --- | --- |
| News channels | 352 | 46.1 | 229 | 43.2 | 114 | 52.8 |
| Public Health and government website | 96 | 12.6 | 66 | 12.5 | 30 | 13.9 |
| Diabetes UK website | 113 | 14.8 | 83 | 15.7 | 28 | 13.0 |
| NHS website | 51 | 6.7 | 35 | 6.6 | 15 | 6.9 |
| Other website | 11 | 1.4 | 7 | 1.3 | 3 | 1.4 |
| Physical/social media | 30 | 3.9 | 26 | 4.9 | 3 | 1.4 |
| Facebook | 31 | 4.1 | 24 | 4.5 | 7 | 3.2 |
| GP, diabetes nurse, healthcare professional | 33 | 4.3 | 22 | 4.2 | 8 | 3.7 |
| Family | 22 | 2.9 | 17 | 3.2 | 4 | 1.9 |
| Friends | 3 | 0.4 | 2 | 0.4 | 1 | 0.5 |
| Employer | 13 | 1.7 | 11 | 2.1 | 2 | 0.9 |
| Diabetes support group | 3 | 0.4 | 3 | 0.6 | 0 | 0.0 |
| Other | 5 | 0.7 | 4 | 0.8 | 1 | 0.5 |
| Not applicable | 1 | 0.1 | 1 | 0.2 | 0 | 0.0 |

| **Which of these resources have you been using for guidance on general diabetes self-management since the start of the pandemic? (Tick all that apply)** | **All**  **(n = 713)** | **%** | **Type 1**  **(n = 487)** | **%** | **Type 2**  **(n = 208)** | **%** |
| --- | --- | --- | --- | --- | --- | --- |
| News channels | 101 | 14.2 | 64 | 13.1 | 34 | 16.4 |
| Public Health and government website | 76 | 10.7 | 48 | 9.9 | 24 | 11.5 |
| Diabetes UK website | 347 | 48.7 | 240 | 49.3 | 99 | 47.6 |
| NHS website | 153 | 21.5 | 99 | 20.3 | 53 | 25.5 |
| Other website | 28 | 3.9 | 19 | 3.9 | 8 | 3.9 |
| Twitter | 37 | 5.2 | 33 | 6.8 | 3 | 1.4 |
| Facebook | 63 | 8.8 | 46 | 9.5 | 17 | 8.2 |
| GP, diabetes nurse, healthcare professional | 207 | 29.0 | 148 | 30.4 | 51 | 24.5 |
| Family | 57 | 8.0 | 42 | 8.6 | 14 | 6.7 |
| Friends | 26 | 3.7 | 20 | 4.1 | 6 | 2.9 |
| Employer | 9 | 1.3 | 6 | 1.2 | 3 | 1.4 |
| Diabetes support group | 48 | 6.7 | 43 | 8.8 | 4 | 1.9 |
| Other | 15 | 2.1 | 9 | 1.9 | 6 | 2.9 |

| **Which of these resources do you use to obtain emotional support? (Tick all that apply)** | **All**  **(n = 687)** | **%** | **Type 1**  **(n = 474)** | **%** | **Type 2**  **(n = 196)** | **%** |
| --- | --- | --- | --- | --- | --- | --- |
| Diabetes UK website – online forum | 55 | 8.0 | 31 | 6.5 | 23 | 11.7 |
| Diabetes UK helpline | 15 | 2.2 | 7 | 1.5 | 7 | 3.6 |
| Physical/social media communities | 71 | 10.3 | 54 | 11.4 | 16 | 8.2 |
| GP, diabetes nurse, healthcare professional | 86 | 12.5 | 61 | 12.9 | 23 | 11.7 |
| Family | 473 | 68.9 | 326 | 68.8 | 134 | 68.4 |
| Friends | 350 | 51.0 | 256 | 54.0 | 86 | 43.9 |
| Employer | 39 | 5.7 | 23 | 4.9 | 16 | 8.2 |
| Diabetes support group | 32 | 4.7 | 28 | 5.9 | 3 | 1.5 |
| Other | 10 | 1.5 | 8 | 1.7 | 2 | 1.0 |

| **Which means do you use to obtain advice/guidance/support from outside your household? (tick all that apply)** | **All**  **(n = 638*)** | **Type 1**  **(n = 442)** | **Type 2**  **(n = 179)** |
| --- | --- | --- | --- |
| Telephone | 124 (19.4%) | 80 (18.1%) | 43 (24.0%) |
| Computer | 467 (73.2%) | 319 (72.2%) | 133 (74.3%) |
| Mobile phone | 429 (67.2%) | 315 (71.3%) | 102 (57.0%) |
| Someone in the house | 41 (6.4%) | 25 (5.7%) | 15 (8.4%) |
| Other | 3 (0.5%) | 2 (0.5%) | 1 (0.6%) |

*113 people said not applicable

**Questions specific to those respondents living alone**

| **If you are living alone, are you receiving support from people outside your household?** | **All*** | **Type 1** | **Type 2** |
| --- | --- | --- | --- |
| No | 83 (50.6%) | 51 (47.2%) | 30 (58.8%) |
| Yes | 81 (49.4%) | 57 (52.8%) | 21 (41.2%) |
|  | 164 (100.0%) | 108 (100.0%) | 51 (100.0%) |

*588 people said NA

This table is restricted to participants who reported living alone for the question on living circumstances.

| **If yes, who from?** | **All**  **(n = 81)** |
| --- | --- |
| Family | 55 (67.9%) |
| Friends | 54 (66.7%) |
| Neighbours | 22 (27.2%) |
| Other | 4 (4.9%) |

**Opinions on information, advice, and support received**

| **In general, how difficult or easy has it been for you to obtain INFORMATION/ADVICE applicable to you on the following?** | **All** | **Type 1** | **Type 2** |
| --- | --- | --- | --- |
| **Glucose control** |  |  |  |
| Very difficult | 34 (5.5%) | 15 (3.5%) | 17 (10.2%) |
| Difficult | 89 (14.4%) | 52 (12.0%) | 34 (20.4%) |
| Moderate | 168 (27.3%) | 123 (28.3%) | 41 (24.6%) |
| Easy | 164 (26.6%) | 125 (28.8%) | 34 (20.4%) |
| Very easy | 161 (26.1%) | 119 (27.4%) | 41 (24.6%) |
| **Diet** |  |  |  |
| Very difficult | 39 (6.2%) | 18 (4.3%) | 21 (10.7%) |
| Difficult | 80 (12.8%) | 46 (11.0%) | 32 (16.3%) |
| Moderate | 174 (27.8%) | 121 (29.0%) | 51 (26.0%) |
| Easy | 173 (27.6%) | 120 (28.8%) | 45 (23.0%) |
| Very easy | 160 (25.6%) | 112 (26.9%) | 47 (24.0%) |
| **Physical activity** |  |  |  |
| Very difficult | 47 (7.3%) | 28 (6.4%) | 19 (9.8%) |
| Difficult | 85 (13.2%) | 55 (12.6%) | 27 (13.9%) |
| Moderate | 163 (25.3%) | 109 (25.1%) | 50 (25.8%) |
| Easy | 199 (30.9%) | 133 (30.6%) | 60 (30.9%) |
| Very easy | 149 (23.2%) | 110 (25.3%) | 38 (19.6%) |
| **Medication** |  |  |  |
| Very difficult | 34 (5.2%) | 21 (4.7%) | 11 (5.8%) |
| Difficult | 94 (14.4%) | 57 (12.8%) | 35 (18.3%) |
| Moderate | 165 (25.3%) | 110 (24.6%) | 50 (26.2%) |
| Easy | 193 (29.6%) | 139 (31.1%) | 51 (26.7%) |
| Very easy | 167 (25.6%) | 120 (26.8%) | 44 (23.0%) |
| **Emotional wellbeing** |  |  |  |
| Very difficult | 84 (13.3%) | 58 (13.5%) | 25 (13.3%) |
| Difficult | 132 (20.9%) | 89 (20.7%) | 40 (21.3%) |
| Moderate | 197 (31.2%) | 136 (31.7%) | 58 (30.9%) |
| Easy | 122 (19.3%) | 79 (18.4%) | 37 (19.7%) |
| Very easy | 96 (15.2%) | 67 (15.6%) | 28 (14.9%) |
| **Diabetes management (if showing symptoms)** |  |  |  |
| Very difficult | 59 (15.7%) | 40 (14.5%) | 19 (20.2%) |
| Difficult | 70 (18.7%) | 47 (17.1%) | 21 (22.3%) |
| Moderate | 103 (27.5%) | 77 (28.0%) | 26 (27.7%) |
| Easy | 79 (21.1%) | 62 (22.5%) | 15 (16.0%) |
| Very easy | 64 (17.1%) | 49 (17.8%) | 13 (13.8%) |
| **Physical/social/physical distancing** |  |  |  |
| Very difficult | 62 (8.7%) | 48 (9.7%) | 13 (6.5%) |
| Difficult | 104 (14.5%) | 81 (16.3%) | 20 (10.0%) |
| Moderate | 169 (23.6%) | 117 (23.5%) | 48 (23.9%) |
| Easy | 191 (26.7%) | 126 (25.4%) | 59 (29.4%) |
| Very easy | 190 (26.5%) | 125 (25.2%) | 61 (30.3%) |

| **In general, how difficult or easy has it been for you to obtain SUPPORT applicable to you on the following?** | **All** | **Type 1** | **Type 2** |
| --- | --- | --- | --- |
| **Glucose control** |  |  |  |
| Very difficult | 63 (12.0%) | 37 (10.1%) | 23 (15.8%) |
| Difficult | 111 (21.1%) | 69 (18.8%) | 39 (26.7%) |
| Moderate | 141 (26.8%) | 102 (27.7%) | 36 (24.7%) |
| Easy | 113 (21.4%) | 86 (23.4%) | 25 (17.1%) |
| Very easy | 99 (18.8%) | 74 (20.1%) | 23 (15.8%) |
| **Diet** |  |  |  |
| Very difficult | 57 (10.9%) | 32 (9.3%) | 24 (14.1%) |
| Difficult | 109 (20.8%) | 67 (19.5%) | 40 (23.5%) |
| Moderate | 144 (27.5%) | 98 (28.6%) | 42 (24.7%) |
| Easy | 122 (23.3%) | 80 (23.3%) | 40 (23.5%) |
| Very easy | 91 (17.4%) | 66 (19.2%) | 24 (14.1%) |
| **Physical activity** |  |  |  |
| Very difficult | 60 (11.1%) | 40 (11.1%) | 19 (11.3%) |
| Difficult | 109 (20.2%) | 67 (18.7%) | 40 (23.8%) |
| Moderate | 145 (26.9%) | 96 (26.7%) | 44 (26.2%) |
| Easy | 127 (23.6%) | 83 (23.1%) | 41 (24.4%) |
| Very easy | 98 (18.2%) | 73 (20.3%) | 24 (14.3%) |
| **Medication** |  |  |  |
| Very difficult | 55 (9.8%) | 34 (9.0%) | 19 (11.0%) |
| Difficult | 102 (18.1%) | 66 (17.5%) | 35 (20.3%) |
| Moderate | 147 (26.1%) | 94 (24.9%) | 48 (27.9%) |
| Easy | 138 (24.5%) | 100 (26.5%) | 33 (19.2%) |
| Very easy | 122 (21.6%) | 83 (22.0%) | 37 (21.5%) |
| **Emotional wellbeing** |  |  |  |
| Very difficult | 89 (16.0%) | 61 (16.2%) | 26 (15.5%) |
| Difficult | 130 (23.4%) | 96 (25.5%) | 32 (19.0%) |
| Moderate | 166 (29.9%) | 103 (27.4%) | 58 (34.5%) |
| Easy | 97 (17.5%) | 66 (17.6%) | 30 (17.9%) |
| Very easy | 73 (13.2%) | 50 (13.3%) | 22 (13.1%) |
| **Diabetes management (if showing symptoms)** |  |  |  |
| Very difficult | 57 (18.7%) | 40 (18.1%) | 16 (20.0%) |
| Difficult | 58 (19.0%) | 43 (19.5%) | 13 (16.3%) |
| Moderate | 84 (27.5%) | 55 (24.9%) | 29 (36.3%) |
| Easy | 55 (18.0%) | 47 (21.3%) | 8 (10.0%) |
| Very easy | 51 (16.7%) | 36 (16.3%) | 14 (17.5%) |
| **Physical/social distancing** |  |  |  |
| Very difficult | 67 (11.1%) | 47 (11.5%) | 17 (9.5%) |
| Difficult | 101 (16.7%) | 77 (18.8%) | 21 (11.7%) |
| Moderate | 165 (27.4%) | 114 (27.9%) | 48 (26.8%) |
| Easy | 143 (23.7%) | 98 (24.0%) | 41 (22.9%) |
| Very easy | 127 (21.1%) | 73 (17.8%) | 52 (29.1%) |
| **How would you rate the QUALITY of the information/advice/support from the following sources or channels?** | **All** | **Type 1** | **Type 2** |
| **Government** |  |  |  |
| Very poor | 105 (14.5%) | 73 (14.3%) | 28 (14.1%) |
| Poor | 178 (24.5%) | 140 (27.5%) | 35 (17.6%) |
| Average | 209 (28.8%) | 137 (26.9%) | 67 (33.7%) |
| Good | 158 (21.8%) | 109 (21.4%) | 46 (23.1%) |
| Very good | 76 (10.5%) | 50 (9.8%) | 23 (11.6%) |
| **Diabetes UK** |  |  |  |
| Very poor | 14 (2.2%) | 10 (2.2%) | 4 (2.2%) |
| Poor | 34 (5.2%) | 23 (5.0%) | 10 (5.6%) |
| Average | 152 (23.4%) | 110 (24.0%) | 38 (21.3%) |
| Good | 293 (45.1%) | 212 (46.3%) | 76 (42.7%) |
| Very good | 157 (24.2%) | 103 (22.5%) | 50 (28.1%) |
| **Physical/social media** |  |  |  |
| Very poor | 56 (9.0%) | 41 (9.5%) | 15 (8.6%) |
| Poor | 150 (24.1%) | 101 (23.3%) | 43 (24.7%) |
| Average | 221 (35.5%) | 152 (35.1%) | 63 (36.2%) |
| Good | 143 (23.0%) | 101 (23.3%) | 40 (23.0%) |
| Very good | 52 (8.4%) | 38 (8.8%) | 13 (7.5%) |
| **News channels** |  |  |  |
| Very poor | 61 (8.5%) | 41 (8.2%) | 18 (9.0%) |
| Poor | 151 (21.1%) | 111 (22.3%) | 38 (19.1%) |
| Average | 234 (32.7%) | 162 (32.5%) | 65 (32.7%) |
| Good | 214 (29.9%) | 147 (29.5%) | 61 (30.7%) |
| Very good | 55 (7.7%) | 37 (7.4%) | 17 (8.5%) |
| **Friends** |  |  |  |
| Very poor | 29 (4.9%) | 19 (4.5%) | 10 (6.3%) |
| Poor | 72 (12.1%) | 49 (11.6%) | 21 (13.3%) |
| Average | 229 (38.4%) | 160 (38.0%) | 63 (39.9%) |
| Good | 199 (33.4%) | 145 (34.4%) | 46 (29.1%) |
| Very good | 67 (11.2%) | 48 (11.4%) | 18 (11.4%) |
| **Family** |  |  |  |
| Very poor | 23 (3.6%) | 14 (3.1%) | 8 (4.6%) |
| Poor | 63 (9.9%) | 43 (9.7%) | 18 (10.3%) |
| Average | 206 (32.3%) | 146 (32.8%) | 54 (31.0%) |
| Good | 229 (35.9%) | 162 (36.4%) | 60 (34.5%) |
| Very good | 116 (18.2%) | 80 (18.0%) | 34 (19.5%) |
| **Employer** |  |  |  |
| Very poor | 80 (17.8%) | 56 (16.5%) | 22 (22.4%) |
| Poor | 65 (14.4%) | 48 (14.1%) | 15 (15.3%) |
| Average | 113 (25.1%) | 79 (23.2%) | 30 (30.6%) |
| Good | 121 (26.9%) | 99 (29.1%) | 19 (19.4%) |
| Very good | 71 (15.8%) | 58 (17.1%) | 12 (12.2%) |
| **Healthcare team** |  |  |  |
| Very poor | 108 (18.0%) | 71 (16.9%) | 36 (22.2%) |
| Poor | 115 (19.2%) | 77 (18.3%) | 34 (21.0%) |
| Average | 116 (19.4%) | 83 (19.7%) | 30 (18.5%) |
| Good | 140 (23.4%) | 101 (24.0%) | 35 (21.6%) |
| Very good | 120 (20.0%) | 89 (21.1%) | 27 (16.7%) |

| Nation/Region of England | n | Healthcare rating good or very good | % |
| --- | --- | --- | --- |
| Scotland | 65 | 26 | 40.0 |
| Wales | 28 | 13 | 46.4 |
| East England | 45 | 20 | 44.4 |
| East Midlands | 32 | 13 | 40.6 |
| Greater London | 47 | 20 | 42.6 |
| North East | 19 | 4 | 21.1 |
| North West | 51 | 21 | 41.2 |
| Northern Ireland | 13 | 5 | 38.5 |
| South East | 132 | 68 | 51.5 |
| South West | 114 | 55 | 48.3 |
| West Midlands | 47 | 14 | 29.8 |

Qualitative responses regarding respondents’ views on way to improve the information, advice and support from the sources above are summarised in the manuscript appendices.

**Personal support networks**

| **For the next questions, please consider your current network of family, friends, contacts.** | **All** | **Living with others** | **Living alone** |
| --- | --- | --- | --- |
| **How would you rate their understanding of your CURRENT diabetes self-management needs?** |  |  |  |
| Very poor | 34 (4.5%) | 25 (3.9%) | 9 (7.9%) |
| Poor | 98 (13.0%) | 82 (12.8%) | 16 (14.0%) |
| Average | 243 (32.1%) | 198 (30.8%) | 45 (39.5%) |
| Good | 225 (29.7%) | 196 (30.5%) | 29 (25.4%) |
| Very good | 157 (20.7%) | 142 (22.1%) | 15 (13.2%) |
| **How would you rate their support in your diabetes self-management during the pandemic?** |  |  |  |
| Very poor | 37 (5.1%) | 29 (4.6%) | 8 (7.5%) |
| Poor | 76 (10.4%) | 64 (10.2%) | 12 (11.2%) |
| Average | 181 (24.7%) | 148 (23.6%) | 33 (30.8%) |
| Good | 240 (32.7%) | 204 (32.6%) | 36 (33.6%) |
| Very good | 199 (27.2%) | 181 (28.9%) | 18 (16.8%) |

| **How has the support you are receiving changed since before the pandemic? (0 = stayed the same)** | **All** | **Type 1** | **Type 2** |
| --- | --- | --- | --- |
| Decreased | 144 (18.8%) | 92 (17.4%) | 49 (22.6%) |
| Same | 405 (53.0%) | 284 (53.7%) | 116 (53.5%) |
| Increased | 215 (28.1%) | 153 (28.9%) | 52 (24.0%) |
|  | 764 (100.0%) | 529 (100.0%) | 217 (100.0%) |

Note: score given on a Likert scale ranging from -5 (Decreased) to 5 (Increased).

| **In what ways do they CURRENTLY support your diabetes self-management? (Tick all that apply)** | **All**  **(n = 603*)** | **%** | **Type 1**  **(n = 309)** | **%** | **Type 2**  **(n = 119)** | **%** |
| --- | --- | --- | --- | --- | --- | --- |
| Food shopping/preparation | 305 | 50.6 | 212 | 50.1 | 85 | 52.2 |
| Picking up medication | 292 | 48.4 | 215 | 50.8 | 69 | 42.3 |
| Essential travel | 84 | 13.9 | 61 | 14.4 | 19 | 11.7 |
| Monitoring blood glucose | 82 | 13.6 | 59 | 14.0 | 21 | 12.9 |
| Emotional support | 374 | 62.0 | 267 | 63.1 | 98 | 60.1 |
| Access to online resources | 65 | 10.8 | 42 | 9.9 | 21 | 12.9 |
| Prompting self-management behaviours | 174 | 28.9 | 121 | 28.6 | 48 | 29.5 |
| Other | 26 | 4.3 | 17 | 4.0 | 4 | 2.5 |

* 141 people said not applicable

### 2. **Responses from parents, carers and partners of people living with diabetes**

#### 2.1. Geographical distribution of responses

| **Region/nation** | **n** | **%** |
| --- | --- | --- |
| Scotland | 24 | 30.4 |
| Wales | 1 | 1.3 |
| Channel Islands | 1 | 1.3 |
| East Midlands | 1 | 1.3 |
| East of England | 6 | 7.6 |
| Greater London | 7 | 8.9 |
| North East | 4 | 5.1 |
| North West | 10 | 12.7 |
| Northern Ireland | 2 | 2.5 |
| South East | 5 | 6.3 |
| South West | 15 | 19.0 |
| West Midlands | 3 | 3.8 |

#### 2.2. Demographic characteristics

|  | **All**  **(n=79)** |
| --- | --- |
| **Gender, n (%)** |  |
| Female | 72 (91%) |
| Male | 7 (9%) |
| **Age, mean (SD)** | 45.2 (10.1) |
| **Ethnicity, n (%)** |  |
| Asian or Asian British: Pakistani | 1 (1%) |
| Black or Black British: Caribbean | 1 (1%) |
| Other Black background | 1 (1%) |
| Other ethnic group | 1 (1%) |
| Other White background | 2 (3%) |
| White: British | 69 (87%) |
| White: Irish | 4 (5%) |

**Living Circumstances**

| **Are you currently living with the person who has diabetes?** | **n (%)** |
| --- | --- |
| No | 7 (9%) |
| Yes | 72 (91%) |
|  | 79 (100%) |

| **Has the number of people you are living with changed as a result of the coronavirus pandemic?** | **n (%)** |
| --- | --- |
| No | 74 (94%) |
| Yes | 5 (6%) |
|  | 79 (100%) |

**Circumstances in relation to COVID-19**

| **Have you been diagnosed with or displayed symptoms of coronavirus since the beginning of February?** | **n (%)** |
| --- | --- |
| No | 67 (85%) |
| Yes | 6 (8%) |
| Not sure | 6 (8%) |
|  | 79 (100%) |

| **Which of the following best describes your current circumstances?** | **n (%)** |
| --- | --- |
| Following stringent Physical/social/physical distancing | 51 (65%) |
| Self-isolating at home | 3 (4%) |
| Shielding group | 4 (5%) |
| Shielding (but not in shielding group) | 0 (0%) |
| Key worker/still leaving home to work | 19 (24%) |
| Minimising interactions to protect someone in household | 2 (3%) |
|  | 79 (100%) |

#### 2.3. Confidence in the ability to support diabetes self-management

| **BEFORE** the coronavirus pandemic and social/physical distancing guidance I was confident that… | **N** | **Median (IQR)** |
| --- | --- | --- |
| Check blood glucose | 64 | 10 (10, 10) |
| Correct high blood glucose | 68 | 10 (8, 10) |
| Correct low blood glucose | 66 | 10 (10, 10) |
| Good blood glucose regulation | 58 | 10 (8, 10) |
| Choose correct foods | 75 | 10 (7, 10) |
| Keep healthy weight | 72 | 8 (5, 10) |
| Examine feet | 78 | 10 (5, 10) |
| Healthy eating pattern | 78 | 8 (6, 10) |
| Physical activity | 78 | 8 (6, 10) |
| Mental wellbeing | 78 | 8 (6, 9) |

Note: score given on a Likert scale ranging from 0 (Could not do at all) to 10 (Certain could do). Not applicable was also an option to account inter-individual variability in condition and self-management requirements.

| **AT PRESENT**, I am confident that… | **N** | **Median (IQR)** |
| --- | --- | --- |
| Check blood glucose | 68 | 10 (10, 10) |
| Correct high blood glucose | 70 | 10 (8, 10) |
| Correct low blood glucose | 67 | 10 (9, 10) |
| Good blood glucose regulation | 57 | 10 (8, 10) |
| Choose correct foods | 75 | 10 (8, 10) |
| Keep healthy weight | 72 | 8 (5, 10) |
| Examine feet | 78 | 10 (5, 10) |
| Healthy eating pattern | 78 | 8 (6, 10) |
| Physical activity | 78 | 8 (4, 10) |
| Mental wellbeing | 78 | 8 (6, 9) |

Note: score given on a Likert scale ranging from 0 (Could not do at all) to 10 (Certain could do). Not applicable was also an option to account inter-individual variability in condition and self-management requirements.

| **Change in score** | **All** |
| --- | --- |
| **Check blood glucose** |  |
| Decreased | 4 (6%) |
| Same | 55 (87%) |
| Increased | 4 (6%) |
| **Correct high blood glucose** |  |
| Decreased | 6 (9%) |
| Same | 50 (76%) |
| Increased | 10 (15%) |
| **Correct low blood glucose** |  |
| Decreased | 9 (14%) |
| Same | 51 (78%) |
| Increased | 5 (8%) |
| **Good blood glucose regulation** |  |
| Decreased | 7 (13%) |
| Same | 38 (68%) |
| Increased | 11 (20%) |
| **Choose correct foods** |  |
| Decreased | 16 (22%) |
| Same | 40 (54%) |
| Increased | 18 (24%) |
| **Keep healthy weight** |  |
| Decreased | 15 (22%) |
| Same | 39 (57%) |
| Increased | 15 (22%) |
| **Examine feet** |  |
| Decreased | 7 (9%) |
| Same | 63 (82%) |
| Increased | 7 (9%) |
| **Healthy eating pattern** |  |
| Decreased | 20 (26%) |
| Same | 45 (58%) |
| Increased | 12 (16%) |
| **Physical activity** |  |
| Decreased | 21 (27%) |
| Same | 49 (64%) |
| Increased | 7 (9%) |
| **Mental wellbeing** |  |
| Decreased | 20 (26%) |
| Same | 48 (62%) |
| Increased | 9 (12%) |

#### 2.4. Sources used for information, advice and support

| **Which of these resources have you used for guidance on how you should behave regarding social/physical distancing measures in relation to the person with diabetes? (Tick all that apply)** | **n**  **(All = 78)** | **%** |
| --- | --- | --- |
| News channels | 47 | 60 |
| Public Health and government website | 33 | 42 |
| Diabetes UK website | 47 | 60 |
| NHS website | 47 | 60 |
| Other website | 3 | 4 |
| Twitter | 8 | 10 |
| Facebook | 19 | 24 |
| GP, diabetes nurse, healthcare professional | 31 | 40 |
| Family | 19 | 24 |
| Friends | 8 | 10 |
| Employer | 5 | 6 |
| Diabetes support group | 12 | 15 |
| Other | 5 | 6 |

| **Which one have you use the most? (Tick one)** | **n** | **%** |
| --- | --- | --- |
| News channels | 26 | 34 |
| Public Health and government website | 9 | 12 |
| Diabetes UK website | 8 | 10 |
| NHS website | 10 | 13 |
| Other website | 0 | 0 |
| Twitter | 2 | 3 |
| Facebook | 4 | 5 |
| GP, diabetes nurse, healthcare professional | 9 | 12 |
| Family | 3 | 4 |
| Friends | 0 | 0 |
| Employer | 1 | 1 |
| Diabetes support group | 4 | 5 |
| Other | 0 | 0 |
|  | 77 | 100 |

| **Which of these resources have you been using for guidance on general diabetes management since the start of the pandemic? (Tick all that apply)** | **n**  **(All = 75)** | **%** |
| --- | --- | --- |
| News channels | 19 | 25 |
| Public Health and government website | 11 | 15 |
| Diabetes UK website | 37 | 49 |
| NHS website | 26 | 35 |
| Other website | 3 | 4 |
| Twitter | 2 | 3 |
| Facebook | 12 | 16 |
| GP, diabetes nurse, healthcare professional | 24 | 32 |
| Family | 5 | 7 |
| Friends | 1 | 1 |
| Employer | 1 | 1 |
| Diabetes support group | 11 | 15 |
| Other | 1 | 1 |

| **Which of these resources do you use to obtain emotional support? (Tick all that apply)** | **n**  **(All = 70)** | **%** |
| --- | --- | --- |
| Diabetes UK website – online forum | 6 | 9 |
| Diabetes UK helpline | 1 | 1 |
| Facebook groups | 12 | 17 |
| GP, diabetes nurse, healthcare professional | 12 | 17 |
| Family | 42 | 60 |
| Friends | 25 | 36 |
| Neighbour | 0 | 0 |
| Employer | 1 | 1 |
| Diabetes support group | 7 | 10 |
| Other | 2 | 3 |

| **Which means do you use to obtain advice/guidance/support from outside your household? (tick all that apply)** | **All**  **(n = 70*)** | **%** |
| --- | --- | --- |
| Telephone | 23 | 33 |
| Computer | 39 | 56 |
| Mobile phone | 50 | 71 |
| Someone in the house | 8 | 11 |
| Other | 0 | 0 |

*7 people said not applicable

#### 2.5. Opinions on information, advice, and support received

| **In general, how difficult or easy has it been for you to obtain INFORMATION/ADVICE applicable to the person you are helping on the following?** | **n (%)** |
| --- | --- |
| **Glucose control** |  |
| Very difficult | 4 (6%) |
| Difficult | 12 (18%) |
| Moderate | 17 (25%) |
| Easy | 21 (31%) |
| Very easy | 14 (21%) |
| **Diet** |  |
| Very difficult | 4 (6%) |
| Difficult | 13 (19%) |
| Moderate | 11 (16%) |
| Easy | 24 (35%) |
| Very easy | 16 (24%) |
| **Physical activity** |  |
| Very difficult | 2 (3%) |
| Difficult | 13 (19%) |
| Moderate | 14 (20%) |
| Easy | 23 (33%) |
| Very easy | 17 (25%) |
| **Medication** |  |
| Very difficult | 5 (7%) |
| Difficult | 13 (18%) |
| Moderate | 16 (23%) |
| Easy | 22 (31%) |
| Very easy | 15 (21%) |
| **Emotional wellbeing** |  |
| Very difficult | 12 (17%) |
| Difficult | 12 (17%) |
| Moderate | 19 (27%) |
| Easy | 15 (21%) |
| Very easy | 13 (18%) |
| **Diabetes management (if showing symptoms)** |  |
| Very difficult | 8 (21%) |
| Difficult | 10 (26%) |
| Moderate | 9 (24%) |
| Easy | 3 (8%) |
| Very easy | 8 (21%) |
| **Physical/social distancing** |  |
| Very difficult | 10 (14%) |
| Difficult | 9 (12%) |
| Moderate | 16 (22%) |
| Easy | 23 (31%) |
| Very easy | 16 (22%) |

| **In general, how difficult or easy has it been for you to obtain SUPPORT applicable to the person you are helping on the following?** | **n (%)** |
| --- | --- |
| **Glucose control** |  |
| Very difficult | 8 (13%) |
| Difficult | 10 (16%) |
| Moderate | 12 (19%) |
| Easy | 16 (26%) |
| Very easy | 16 (26%) |
| **Diet** |  |
| Very difficult | 7 (11%) |
| Difficult | 7 (11%) |
| Moderate | 15 (24%) |
| Easy | 19 (30%) |
| Very easy | 15 (24%) |
| **Physical activity** |  |
| Very difficult | 6 (10%) |
| Difficult | 10 (16%) |
| Moderate | 13 (21%) |
| Easy | 21 (33%) |
| Very easy | 13 (21%) |
| **Medication** |  |
| Very difficult | 7 (11%) |
| Difficult | 7 (11%) |
| Moderate | 13 (20%) |
| Easy | 20 (31%) |
| Very easy | 17 (27%) |
| **Emotional wellbeing** |  |
| Very difficult | 16 (25%) |
| Difficult | 6 (9%) |
| Moderate | 17 (26%) |
| Easy | 13 (20%) |
| Very easy | 13 (20%) |
| **Diabetes management (if showing symptoms)** |  |
| Very difficult | 9 (26%) |
| Difficult | 5 (15%) |
| Moderate | 8 (24%) |
| Easy | 4 (12%) |
| Very easy | 8 (24%) |
| **Physical/social distancing** |  |
| Very difficult | 10 (16%) |
| Difficult | 10 (16%) |
| Moderate | 13 (20%) |
| Easy | 16 (25%) |
| Very easy | 15 (23%) |

| **How would you rate the QUALITY of the information/advice/support from the following sources or channels?** | **n (%)** |
| --- | --- |
| **Government** |  |
| Very poor | 11 (14%) |
| Poor | 15 (19%) |
| Average | 23 (30%) |
| Good | 19 (25%) |
| Very good | 9 (12%) |
| **Diabetes UK** |  |
| Very poor | 1 (1%) |
| Poor | 3 (4%) |
| Average | 13 (19%) |
| Good | 29 (42%) |
| Very good | 23 (33%) |
| **Physical/social media** |  |
| Very poor | 7 (10%) |
| Poor | 9 (13%) |
| Average | 29 (43%) |
| Good | 18 (26%) |
| Very good | 5 (7%) |
| **News channels** |  |
| Very poor | 8 (11%) |
| Poor | 16 (23%) |
| Average | 24 (34%) |
| Good | 14 (20%) |
| Very good | 9 (13%) |
| **Friends** |  |
| Very poor | 4 (7%) |
| Poor | 7 (12%) |
| Average | 22 (38%) |
| Good | 19 (33%) |
| Very good | 6 (10%) |
| **Family** |  |
| Very poor | 4 (6%) |
| Poor | 8 (13%) |
| Average | 22 (34%) |
| Good | 20 (31%) |
| Very good | 10 (16%) |
| **Employer** |  |
| Very poor | 8 (18%) |
| Poor | 10 (23%) |
| Average | 16 (36%) |
| Good | 8 (18%) |
| Very good | 2 (5%) |
| **Healthcare team** |  |
| Very poor | 3 (5%) |
| Poor | 10 (15%) |
| Average | 13 (20%) |
| Good | 14 (22%) |
| Very good | 25 (38%) |

#### 2.6. Own role in supporting someone with diabetes

| **How would you rate your understanding of their CURRENT diabetes self-management needs?** | **n (%)** |
| --- | --- |
| Very poor | 1 (1%) |
| Poor | 3 (4%) |
| Average | 11 (14%) |
| Good | 26 (33%) |
| Very good | 37 (47%) |

| **In what ways do you CURRENTLY support the individual in their diabetes self-management? (Tick all that apply)** | **n = 78*** | **%** |
| --- | --- | --- |
| Food shopping/preparation | 69 | 88 |
| Picking up medication | 67 | 86 |
| Essential travel | 49 | 63 |
| Monitoring blood glucose | 49 | 63 |
| Emotional support | 69 | 88 |
| Access to online resources | 41 | 53 |
| Prompting self-management behaviours | 69 | 88 |
| Other | 6 | 8 |

*1 person said not applicable
